# Bexmarilimab-induced macrophage activation leads to treatment benefit in solid tumors: the phase I/II first-in-human MATINS trial

**DOI:** 10.1101/2023.04.17.23288693

**Authors:** Jenna H. Rannikko, Loic Verlingue, Maria de Miguel, Annika Pasanen, Debbie Robbrecht, Tanja Skytta, Sanna Iivanainen, Shishir Shetty, Yuk Ting Ma, Donna M. Graham, Sukeshi Patel Arora, Panu Jaakkola, Christina Yap, Yujuan Xiang, Jami Mandelin, Matti K. Karvonen, Juho Jalkanen, Sinem Karaman, Jussi P. Koivunen, Anna Minchom, Maija Hollmén, Petri Bono

**Author notes:** Corresponding authors: Maija Hollmén, PhD, MediCity Research Laboratory, University of Turku, Tykistökatu 6A, 20520 Turku, Finland,; Petri Bono, MD, PhD, Terveystalo Finland and University of Helsinki, Jaakonkatu 3B, 00100 Helsinki, Finland.

## Abstract

Clever-1 expression in macrophages contributes to impaired antigen presentation and suppression of anti-tumor immunity. This first-in-human trial was designed to investigate the safety and tolerability of Clever-1 blockade in patients with treatment-refractory solid tumors and to assess preliminary anti-tumor efficacy, pharmacodynamics, and immunologic correlates. Bexmarilimab was well tolerated with no observed dose limiting toxicities in part I (n=30) and no additional safety signals in part II (n=108). Disease control (DC) rates of 25-40% were observed in cutaneous melanoma, gastric, hepatocellular, estrogen receptor positive breast, and biliary tract cancers, which associated with improved survival in a landmark analysis. High pre-treatment intra-tumoral Clever-1 staining and low circulating TNFα correlated with DC. Digital spatial profiling of DC and non-DC tumors with next generation sequencing showed bexmarilimab-induced macrophage activation and robust stimulation of IFNγ and T-cell receptor signaling selectively in DC patients. These data suggest that bexmarilimab therapy is well-tolerated and show that macrophage targeting can promote tumor control in late stage cancer.

**Highlights:**

- Targeting Clever-1 with bexmarilimab is well tolerated
- Disease control in late stage cancer can be achieved with bexmarilimab monotherapy
- Low baseline immune activation associates with bexmarilimab benefit
- Bexmarilimab converts intratumoral macrophages to support adaptive immune responses

## Introduction

In recent years, immune checkpoint blockade has become a standard of care approach in numerous malignancies. Anti-PD-(L)1 agents inhibit T-cell checkpoints resulting in T-cell activation and cancer cell killing. However, primary and secondary resistance is common, and attempts have been made to overcome this with patient selection, combinatory approaches, and new immunotherapeutic treatment modalities.

Mounting evidence implicates the tumor microenvironment (TME) in driving resistance to immune checkpoint therapy^1^. The accumulation of myeloid cells in the TME has been shown to promote tumor growth in several pre-clinical models^2^. Clever-1 is a multifunctional scavenger and adhesion receptor expressed by human monocytes, subsets of immunosuppressive macrophages (M2 type), lymphatic endothelial cells, and sinusoidal endothelial cells^3, 4^. In macrophages, Clever-1 is involved in receptor-mediated endocytosis and recycling, intracellular sorting and transcytosis of altered and normal self-components. In several cancers, high Clever-1 expression is associated with poor prognosis, T-cell exclusion, impaired antigen presentation, and resistance to immune checkpoint inhibitors^5–7^. Targeting Clever-1 in various *in vivo* models delays tumor growth by activating cytotoxic CD8+ T-cells and improves responsiveness to anti-PD-1 therapy in refractory models^8, 9^.

Bexmarilimab (FP-1305) is a humanized monoclonal IgG4 antibody specific for human Clever-1. It contains IgG4 (S241P) heavy-chain and kappa light-chain constant regions and has been further optimized by introducing the L248E mutation to avoid Fc receptor binding^10^. Thus, bexmarilimab has very low antibody-dependent cell cytotoxicity and complement-mediated effector functions. Functionally, bexmarilimab potentiates TNFα release after LPS stimulus^10^ and impairs multiprotein vacuolar ATPase-mediated acidification of phago-lysosomes and leads to improved antigen presentation of scavenged antigens in primary human macrophages^11^.

To investigate the potential of bexmarilimab in inducing anti-tumor immune responses a phase I/II first-in-human clinical trial (MATINS; NCT03733990) was designed to study the safety, tolerability and early efficacy of bexmarilimab in patients with selected advanced or metastatic solid tumors (Supplementary Fig. S1). This basket trial approach enabled us to identify responding cancer types and biomarkers related to response. Indeed, initial results from part I of the MATINS trial shows evidence that targeting Clever-1 promotes peripheral T-cell activation in a subset of patients^11^. Here we report part I and part II of the MATINS trial and show that patients responding favorably to bexmarilimab have low baseline systemic cytokine levels. With digital spatial profiling of pre- and post-tumor biopsy samples we show that the response coincided with intratumoral macrophage conversion and induction of adaptive immune responses.

## Results

### Patients and Treatments in part I

Between 13 Dec 2018 - 16 Jan 2020, 30 patients with advanced colon, pancreatic, biliary tract, ovarian or hepatocellular carcinomas, or checkpoint-inhibitor refractory cutaneous melanomas commenced intravenous bexmarilimab in five escalating dose cohorts of 0.1, 0.3, 1, 3, and 10 mg/kg Q3W with five to seven patients in each dose level. The median age of included patients was 65 (range, 30-81) and all had exhausted standard-of-care treatment options with a median number of four treatment lines for advanced disease (**Table 1**; range, 1-8). The median number of doses for bexmarilimab was three to five across the different dose cohorts and the median time on treatment was 2.2 months (range, 0.8 to 13.7).

**Table 1.** Patient characteristics

| **Characteristic** |  | **Number (%)**<br>**Part I**<br>n=30 | **Number (%)**<br>**Part II**<br>n=108 |
| --- | --- | --- | --- |
| **Age, median** |  | 65 | 60 |
| **Gender** |  |  |  |
|  | Male | 8 (27) | 59 (54) |
|  | Female | 22 (73) | 51 (46) |
| **ECOG PS** |  |  |  |
|  | 0 | 11 (37) | 45 (41) |
|  | 1 | 19 (63) | 65 (59) |
| **Cancer type** |  |  |  |
|  | Colorectal cancer | 15 (50) | 28*(26) |
|  | Pancreatic cancer | 8 (27) | 10 (9) |
|  | Ovarian cancer | 2 (7) | 10 (9) |
|  | Cutaneous melanoma | 2 (7) | 10 (9) |
|  | Biliary tract cancer | 2 (7) | 10 (9) |
|  | Hepatocellular carcinoma | 1 (3) | 10 (9) |
|  | Gastric adenocarcinoma | NA | 10 (9) |
|  | Uveal melanoma | NA | 10 (9) |
|  | ER+ breast cancer | NA | 10 (9) |
| **No of lines, previous therapies (median)** |  | 4 | 3 |
| **PD-1/PD-L1 antibodies as prior therapy** |  |  |  |
|  | Cutaneous melanoma | 2 (100) | 10 (100) |
|  | Hepatocellular carcinoma | 0 (0) | 4 (40) |
|  | Colorectal cancer | 0 (0) | 2 (7) |
|  | Ovarian cancer | 1 (50) | 2 (20) |
|  | Uveal melanoma | NA | 1 (10) |

ECOG PS: Eastern Cooperative Oncology Group performance status; \*Two part I patients were included in part II

### Pharmacokinetics, Pharmacodynamics and Immunogenicity in part I

The collected blood samples for determination of bexmarilimab showed rapid elimination of the investigational medicinal product (IMP) with a terminal half-life of 13.9 hours and a dose proportional elimination (Supplementary Fig. S2A). Over 50% receptor occupancy in circulating monocytes was observed at day 2 with dose levels higher than 1mg/kg (Supplementary Fig. S2B), but this level of occupancy did not remain throughout the entire treatment cycle and was markedly reduced by day 8 (Supplementary Fig. S2B). Longer-lasting neutralization of circulating Clever-1 up to -70% from baseline was observed at higher doses (3-10mg/kg) and resulted in marked downregulation lasting for 8-15 days (Supplementary Fig. S2C). Anti-drug antibodies were observed in three patients (12%) at cycle 2. As reported earlier, all tested dose levels resulted in increased circulating NK and CD8+ T-cells, B-cells and decreased number of circulating FoxP3+ regulatory T-cells. Furthermore, circulating CD8+ and CD4+ cells showed upregulation of CD25, CXCR3 and CD69 suggesting activation of adaptive immune responses ^11^.

### Patients and Treatments in part II

Based on the clinical efficacy and circulating biomarker data seen in part I of the study, the data monitoring committee of the trial recommended a dose selection of 1.0 mg/kg Q3W for part II and additional cohorts of 0.3 mg/kg and 3 mg/kg Q3W in colorectal cancer. For part II, the patients commenced treatment between 22 Feb 2020 – 16 Apr 2021. At the data cut-off, 108 patients dosed in eleven cohorts (ten patients per cohort) of specific cancer types were fully enrolled while the ATC cohort is still recruiting patients. Patient characteristics are presented in **Table 1**. In brief, the patients had received a median of three previous therapy lines in the advanced disease setting (range, 0 to 8).

Previous PD-(L)1 targeted therapy was used in all cutaneous melanoma (100%), four HCC (40%), two ovarian cancer (20%) and two CRC patients (7%).

### Safety in part I and part II

In part I of the study, no dose-related toxicities (DLT) were observed, and maximum tolerated dose (MTD) could not be defined. In part I and II, 138 patients who had received at least one dose of bexmarilimab, 50% reported treatment-related adverse events (TRAE), 8.7% of them were grade 3- 4 events, and no treatment-related deaths were observed. Of the TRAEs, fatigue, pyrexia, and anemia were the most common and predominantly presented as low-grade events (**Table 2**). No association between the dose of bexmarilimab and TRAE was observed (Supplementary Table S2). Of the grade ≥3 treatment-emergent adverse events (TEAE), anemia (5.8%), and ascites (5.8%) were the most common (Supplementary Table S3). Since the proposed main mode of action of bexmarilimab is immune activation, immune-related adverse events were of special interest. Immune-related adverse events (dermatitis, myositis, thyroiditis, pneumonitis of grade 2 and hepatitis of grade 4) leading to treatment discontinuation were recorded in two patients in part I cycles 8 and 11, respectively. The observed immune-related adverse events responded to bexmarilimab discontinuation and administration of systemic corticosteroids. In part II, five patients developed immune-mediated adverse events of grade ≥2 (hyperthyroidism, rash, pneumonitis, pancreatic insufficiency, hepatitis), all of which resolved without sequelae.

**Table 2.**
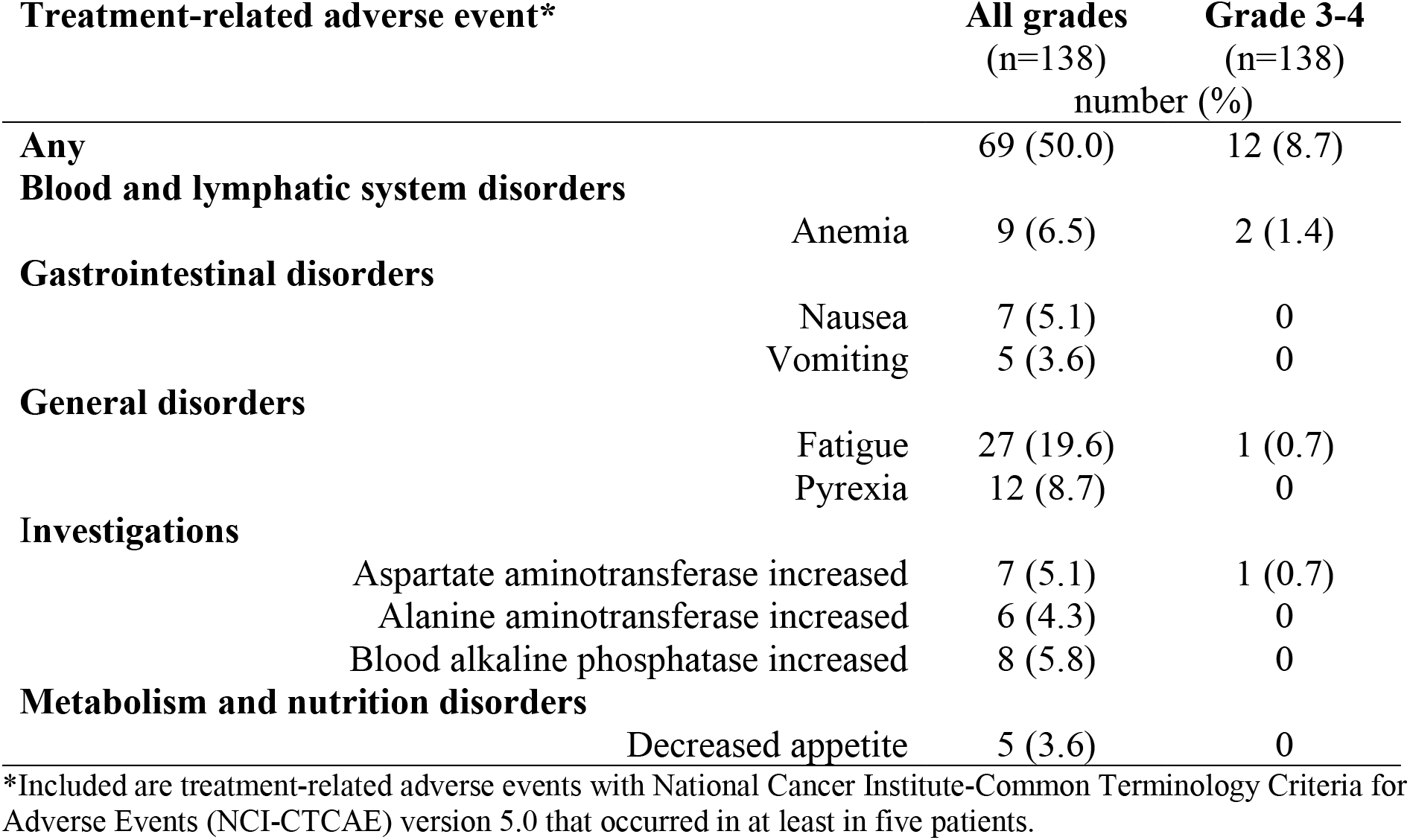
Treatment-related adverse events part I and part II

| Treatment-related adverse event* |  | All grades<br>(n=138)<br>number (%) | Grade 3-4<br>(n=138)<br>number (%) |
| --- | --- | --- | --- |
| **Any** |  | 69 (50.0) | 12 (8.7) |
| **Blood and lymphatic system disorders** |  |  |  |
|  | Anemia | 9 (6.5) | 2 (1.4) |
| **Gastrointestinal disorders** |  |  |  |
|  | Nausea | 7 (5.1) | 0 |
|  | Vomiting | 5 (3.6) | 0 |
| **General disorders** |  |  |  |
|  | Fatigue | 27 (19.6) | 1 (0.7) |
|  | Pyrexia | 12 (8.7) | 0 |
| **Investigations** |  |  |  |
|  | Aspartate aminotransferase increased | 7 (5.1) | 1 (0.7) |
|  | Alanine aminotransferase increased | 6 (4.3) | 0 |
|  | Blood alkaline phosphatase increased | 8 (5.8) | 0 |
| **Metabolism and nutrition disorders** |  |  |  |
|  | Decreased appetite | 5 (3.6) | 0 |

\*Included are treatment-related adverse events with National Cancer Institute-Common Terminology Criteria for Adverse Events (NCI-CTCAE) version 5.0 that occurred in at least in five patients.

### Efficacy in part I and II

In part I of the study, two patients, one with colorectal cancer (CRC) (pretreated with five previous lines of systemic therapy) and one with cutaneous melanoma (pretreated with four previous lines of systemic therapy including anti-PD-1 and CTLA-4) had tumor shrinkage in response to bexmarilimab at dose levels of 0.3 and 1 mg/kg, respectively. The microsatellite stable, K-RAS wild type CRC patient had a long-lasting partial response (PR) according to RECIST 1.1 and the treatment was stopped after cycle eight due to immune-related toxicity while the response lasted until end of follow-up (over 100 days from last dose of bexmarilimab) (Supplementary Fig. S3A and B). The melanoma patient who had PR in target lesions simultaneously developed symptomatic brain metastasis and was taken off study.

Of the patients in part I and II, RECIST 1.1 defined PR rate at cycle 4 was observed in one patient (1%) and stable disease (SD) in 18 patients (13%) corresponding to disease control rate (DCR) of 14%. DCR was the highest in cutaneous melanoma (25%), biliary tract cancers (25%), gastric adenocarcinoma (30%), hepatocellular cancer (40%), and ER+ breast cancer (40%) (**Table 3**), translating to prolonged treatment durations (**Fig. 1A**). Progression-free survival (PFS) and overall survival (OS) were analyzed in all patients treated in part I and II of the study (n=138). The median PFS was 59 days (95% CI, 58-61), and OS 151 days (95% CI, 118-190) (**Fig. 1B**). To define the relationship between disease control (DC) and OS, an exploratory landmark analysis according to response was conducted. The results showed improved survival (HR 0.139, 95% CI 0.034-0.575) for patients with DC compared to non-DC patients at cycle four (**Fig. 1C**) while the duration of the previous line of therapy prior to entering the trial was similar for patients with DC or non-DC (**Fig. 1D**). The survival difference between DC and non-DC patients was the most drastic in cutaneous melanoma and bile duct cancers where no deaths were observed in the DC patients (Supplementary Fig. S4A and B). The PFS ratio (defined by the PFS on bexmarilimab/duration of previous treatment line) was >1.3 in 13% patients. The PFS ratio was positive in 42% of DC and 12% of non-DC patients (p=0.0031), respectively (Supplementary Table S4).

**Figure 1.**
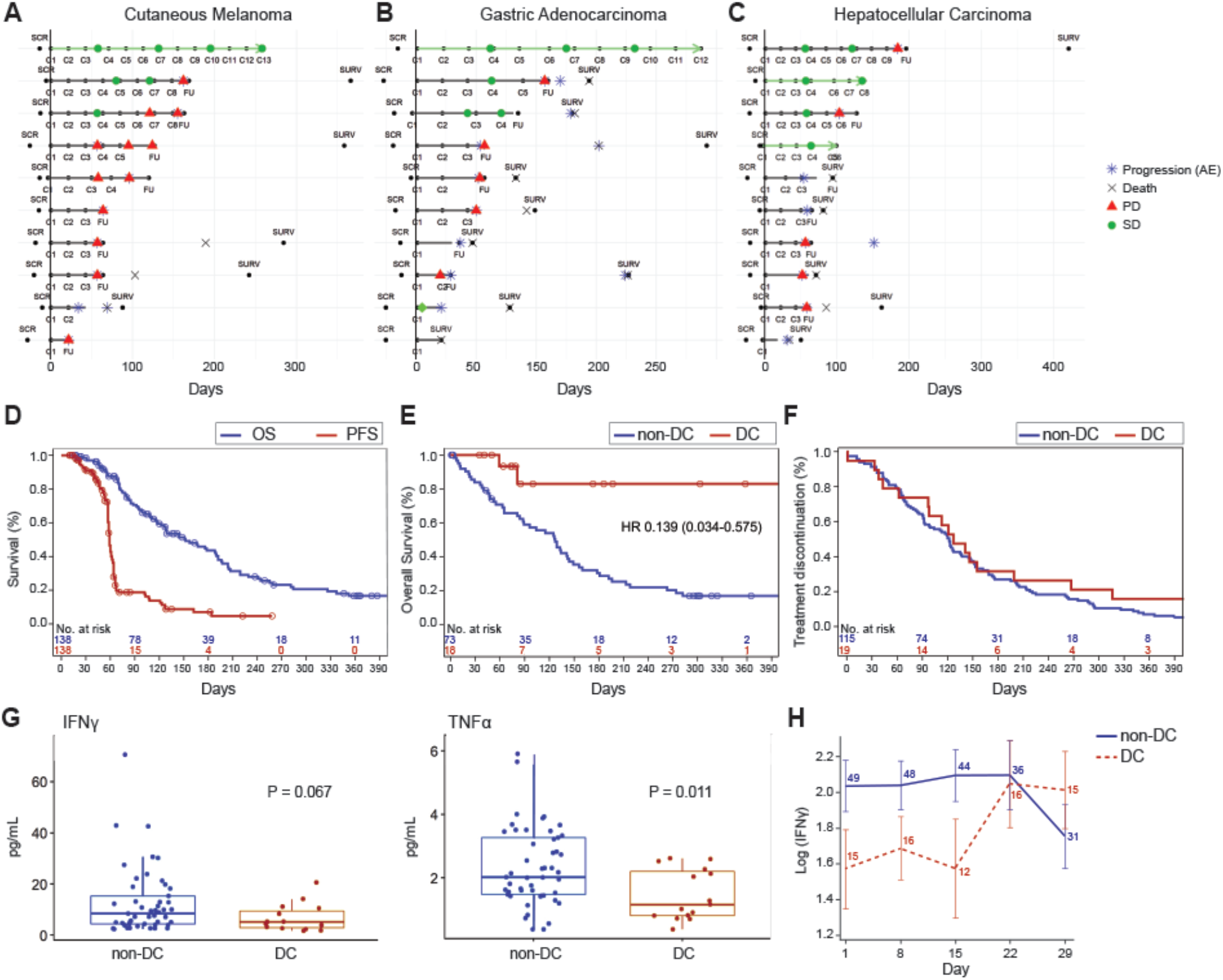
Bexmarilimab responses in patients showing disease control. A, Swimmer-plot analysis for treatment durations and tumor responses in melanoma, gastric and hepatocellular cancer patients of part II. Y-axis shows individual patients, X-axis time in days from the first bexmarilimab dose. SCR: screening time; SURV: survival visit; C: Cycle number; Green circle: SD response; Red triangle: PD-response; Blue asterisk: PD reported as AE; X: death of subject. **B**, Kaplan-Meier analysis for PFS (red) and OS (blue). **C**, Landmark Kaplan-Meier analysis for OS from cycle four according to PR/SD (red) or PD (blue). **D**, Kaplan-Meier analysis for previous therapy line treatment duration (prior entering the trial) according to PR/SD (red) or PD (blue). Circles indicate censored events. Y-axis, time in days. **E**, Baseline levels of IFNγ and TNFα according to DC status. **F**, Change of IFNγ levels during the first cycle of treatment according to DC. Blue: non-DC patients with bexmarilimab; Red: DC patients with bexmarilimab.

**Table 3.** ORR and DC rates in part I and II at cycle 4 in part I and II.

|  |  | Number (%) |
| --- | --- | --- |
|  |  | n=138 |
| **ORR** |  | 1 (1) |
|  | CR | 0 (0) |
|  | PR | 1 (1) |
|  | SD | 18 (13) |
|  | PD | 119 (86) |
| **DCR** |  | 19 (14) |
|  | Colorectal cancer (n=43) | 2 (5) |
|  | Pancreatic cancer (n=18) | 0 (0) |
|  | Ovarian cancer (n=12) | 0 (0) |
|  | Cutaneous Melanoma (n=12) | 3 (25) |
|  | Biliary tract cancer (n=12) | 3 (25) |
|  | Hepatocellular carcinoma (n=11) | 4 (36) |
|  | Gastric adenocarcinoma (n=10) | 3 (30) |
|  | Uveal melanoma (n=10) | 0 (0) |
|  | ER+ breast cancer (n=10) | 4 (40) |

ORR: objective response rate; CR: complete response; PR: partial response; SD: stable disease; PD: progressive disease; DCR: disease control rate

### Biomarker analysis of MATINS patients

Pre-treatment tumor samples were stained for Clever-1 and PD-L1 in the tumor cohorts with the highest DC (cutaneous melanoma, gastric, biliary tract and hepatocellular carcinoma). 78 (100%) and 43 patients (55%) had pre-treatment tumor biopsy staining results available for Clever-1 and PD-L1, respectively. The percentages for Clever-1 positivity and PD-L1 are presented in Table 4. Tumor Clever-1 and PD-L1 scores were compared between the patients with DC and non-DC at cycle four. Higher Clever-1 positivity in the intra-tumor location correlated significantly with DC (median 15% vs 3%, p=0.038). Lower PD-L1 CPS was observed in DC patients (median 1%, range 0-2%) compared to non-DC (median 5%, range 0-100%) but this was not statistically significant (**Table 4**, Supplementary Fig. S5A and B).

**Table 4.**
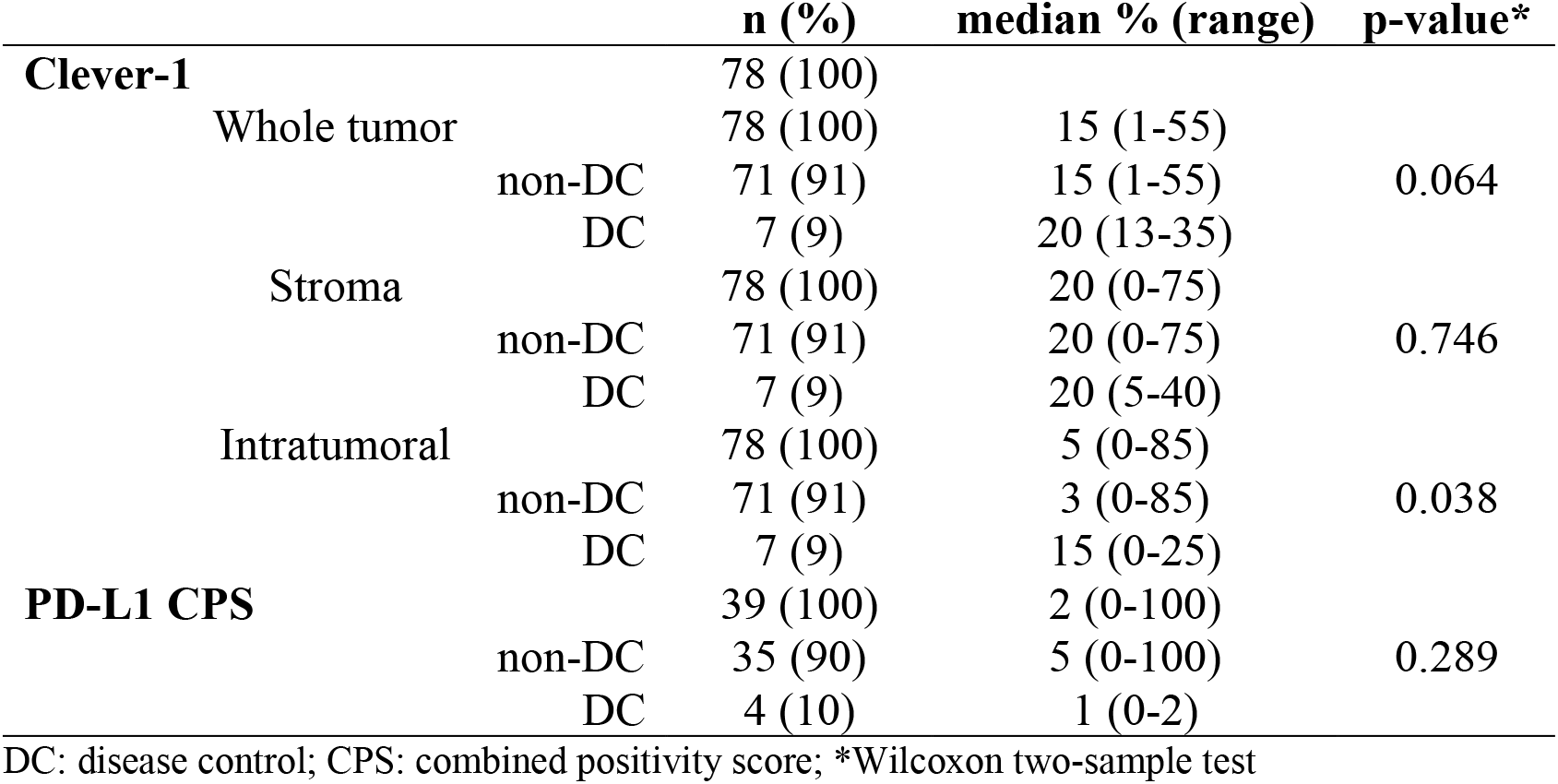
Clever-1 and PD-L1 expression in pre-treatment tumors

Circulating cytokine analysis at baseline and over the treatment were studied in the tumor cohorts with the highest DC rates (n=64). Low baseline TNFα was associated with DC (p=0.0011) and there was tendency for lower IFNγ levels for DC patients (**Fig. 1E**). No change over time in the serum TNFα levels was observed (not shown) while increases of IFNγ levels (day 22-29) were observed with DC patients (**Fig. 1F**).

### Profiling of DC and non-DC patient tumor gene expression by spatial transcriptomics

To more comprehensively study bexmarilimab activity in the TME, we performed GeoMx digital spatial profiling with next generation sequencing on MATINS pre- and post-treatment biopsies. For the analysis, we selected two cancer types with high DC, biliary tract cancer and ER+ breast cancer, and included patients with both pre- and post-treatment biopsies available (DC, n = 3; non-DC, n = 3). The biopsies were stained for CD68, CD31 and pan-cytokeratin to analyze gene expression separately in macrophages (CD68^+^), vessels (CD31^+^) and the remaining tumor area (CD68^-^CD31^-^) (**Fig. 2A**, Supplementary Fig. S6A and Methods). Both CD68^+^ and CD31^+^ subsets are characterized to be Clever-1 positive in various tumors^5^.

**Figure 2.**
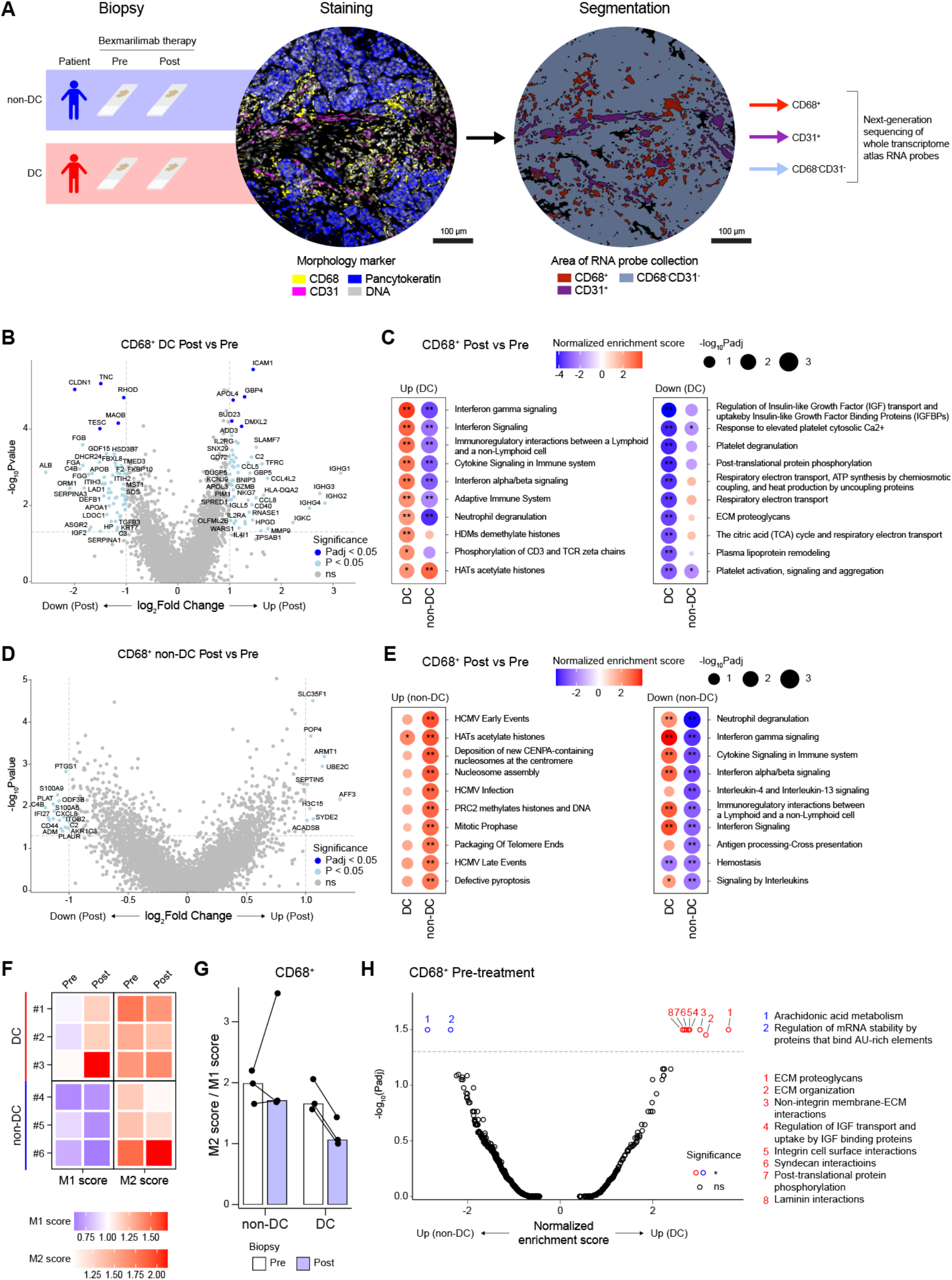
GeoMx spatial profiling reveals pro-inflammatory conversion of tumor-associated macrophages in DC patients. **A**, Schematic for GeoMx spatial transcriptomics profiling of pre- and post-treatment biopsies from non-DC (n = 3 patients, 30 ROIs) and DC (n = 3 patients, 33 ROIs) patients. Representative images of morphology marker staining and corresponding segmentation are displayed for a single ROI. **B**, Volcano plot showing differentially expressed genes in DC patient CD68^+^ areas after bexmarilimab therapy. **C**, Bubble plots of top up- and downregulated pathways in CD68^+^ areas of DC patients after bexmarilimab therapy (gene set enrichment analysis). **D**, Volcano plot showing differentially expressed genes in non-DC patient CD68^+^ areas after bexmarilimab therapy. **E**, Bubble plots of top up- and downregulated pathways in CD68^+^ areas of non-DC patients after bexmarilimab therapy (gene set enrichment analysis). **F**, Heatmap of M1 and M2 macrophage gene scores on CD68^+^ area. Red color indicates higher M1 (or M2) gene expression than overall gene expression, blue color lower. Each tile represents median of patient’s ROIs. **G**, Bar graph of M2 and M1 score ratios corresponding to scores shown in **F**. Each point represents patient median across ROIs and each bar patient group median. **H**, Volcano plot showing differently activated pathways between CD68^+^ areas of non-DC and DC pre-treatment biopsies (gene set enrichment analysis). DC, disease control during bexmarilimab therapy; non-DC, no disease control during bexmarilimab therapy; Pre, pre-treatment biopsy; Post, post-treatment biopsy; ROI, region of interest; **, Padj < 0.05; *, Padj < 0.05; ns, not significant. In **C** and **E**, red color denotes pathway activation and blue downregulation.

Gene expression profiles of 63 biopsy regions, corresponding to a total of 180 segments (63 CD68^+^ and CD68^-^CD31^-^ areas, 54 CD31^+^ areas) were obtained after excluding segments with low signal-to-noise ratio (Supplementary Fig. S6B). The overall gene expression profiles of the segments were compared by unsupervised hierarchical clustering (Supplementary Fig. S6C) and principal component analysis (Supplementary Fig. S6D), which showed that the segment type and cancer type primarily drove differences in gene expression, as expected after successful segmentation. The expression of Clever-1 (*STAB1*) was most abundant in the CD68^+^ and CD31^+^ segments validating cell type specificity of the segmentation thresholds (Supplementary Fig. S7A).

### Bexmarilimab activates interferon signaling and M1-like gene expression in tumor-associated macrophages (TAM) of MATINS DC patients

By investigating the gene expression changes in TAMs (CD68+) between pre-and post-treatment biopsies we observed upregulation of interferon-inducible genes (*ICAM1, SLAMF7*, *GBP4*, *GBP5, CD40*) as well as chemokines (*CCL4L2*, *CCL5, CCL2*) (**Fig. 2B**, Supplementary Data Files) selectively in DC patients. For instance, SLAMF7 drives strong activation and pro-inflammatory cytokine secretion in macrophages under inflammatory conditions^12^. Moreover, significant upregulation of interferon signaling, antigen presentation on class I major histocompatibility complex (MHC) and adaptive immune response related pathways was observed in DC TAMs after bexmarilimab therapy (**Fig. 2C**, Supplementary Fig. S7B and Supplementary Data Files). In strong contrast, non-DC TAMs showed only few differentially expressed genes and downregulation of pathways related to immune system activation, such as interferon gamma signaling, antigen processing and interleukin signaling (**Fig. 2D** and **E**).

To further examine the changes in TAM phenotypes after bexmarilimab therapy, we calculated M1 and M2 macrophage scores in each CD68^+^ segment based on the M1-M2 macrophage genes published by Martinez et al^13^ (Supplementary Fig. S7C, D and E). The M1 scores increased and the M2/M1 score ratios decreased in all DC patients after bexmarilimab therapy, while similar changes were not observed in non-DC patients (**Fig. 2F** and **G**), suggesting a beneficial pro-inflammatory conversion of TAMs specifically in DC patient tumors.

Unlike the robust response observed in the CD68^+^ tumor area, the CD31^+^ vessels showed a lower level of gene expression changes in both DC and non-DC patients. The altered pathways were not directly related to immune system function and rather showed changes in translation and ECM modulation (Supplementary Fig. S8A to D). Notably, the integrin cell surface interaction pathway was downregulated during the treatment in both DC and non-DC vessels indicating decreased leukocyte binding (Supplementary Fig. S8B and D). Indeed, the parent antibody (3-372) of bexmarilimab has been shown to decrease leukocyte binding and transmigration via vascular endothelium^14, 15^.

While investigating whether pre-existing differences in TAM phenotypes could explain bexmarilimab response, we found only a few significantly altered pathways between DC and non-DC patients. Essentially, DC TAMs expressed higher levels of genes modulating the extracellular matrix (**Fig. 2H**). In summary, these data demonstrate the ability of bexmarilimab to activate TAMs within human tumors. Whereas clearly different responses were observed in DC and non-DC TAMs, the TAMs were not widely different prior to therapy.

### TAM activation is reflected to adjacent immune cells

We next evaluated how the CD68^-^CD31^-^ tumor region was altered in DC patients to reflect the observed pro-inflammatory activation of TAMs. Significantly increased transcription was observed for interferon signaling genes (*IFI16, IFI44, GBP1, GBP5, CD40*), immune cell attracting chemokines (*CCL4L2, CCL5, CXCL9, CCL2, CXCL16*), lymphocyte and natural killer (NK) cell markers (*CD3E, CD8A, CD2, NKG7, FCGR3A*) and MHC class I and II proteins (e.g., *HLA-B, HLA-DRA, HLA-DPA1*) (**Fig. 3A** and Supplementary Data Files). These signs of strong immune activation were confirmed by pathway analysis that showed significant upregulation of interferon gamma signaling and phosphorylation of CD3 and TCR zeta chains after bexmarilimab therapy (**Fig. 3B**). As with TAMs, similar changes were not observed in non-DC TME (Supplementary Fig. S8E and F). Using the SpatialDecon algorithm we quantified the immune cell types in the CD68^-^CD31^-^ segment (Supplementary Fig. S8G and Methods), which showed bexmarilimab-induced increase in CD4^+^ and CD8^+^ T-cell abundancies in both DC and non-DC patients, while NK cell, B-cell and macrophage abundancies were increased exclusively in DC patients (**Fig. 3C**).

**Figure 3.**
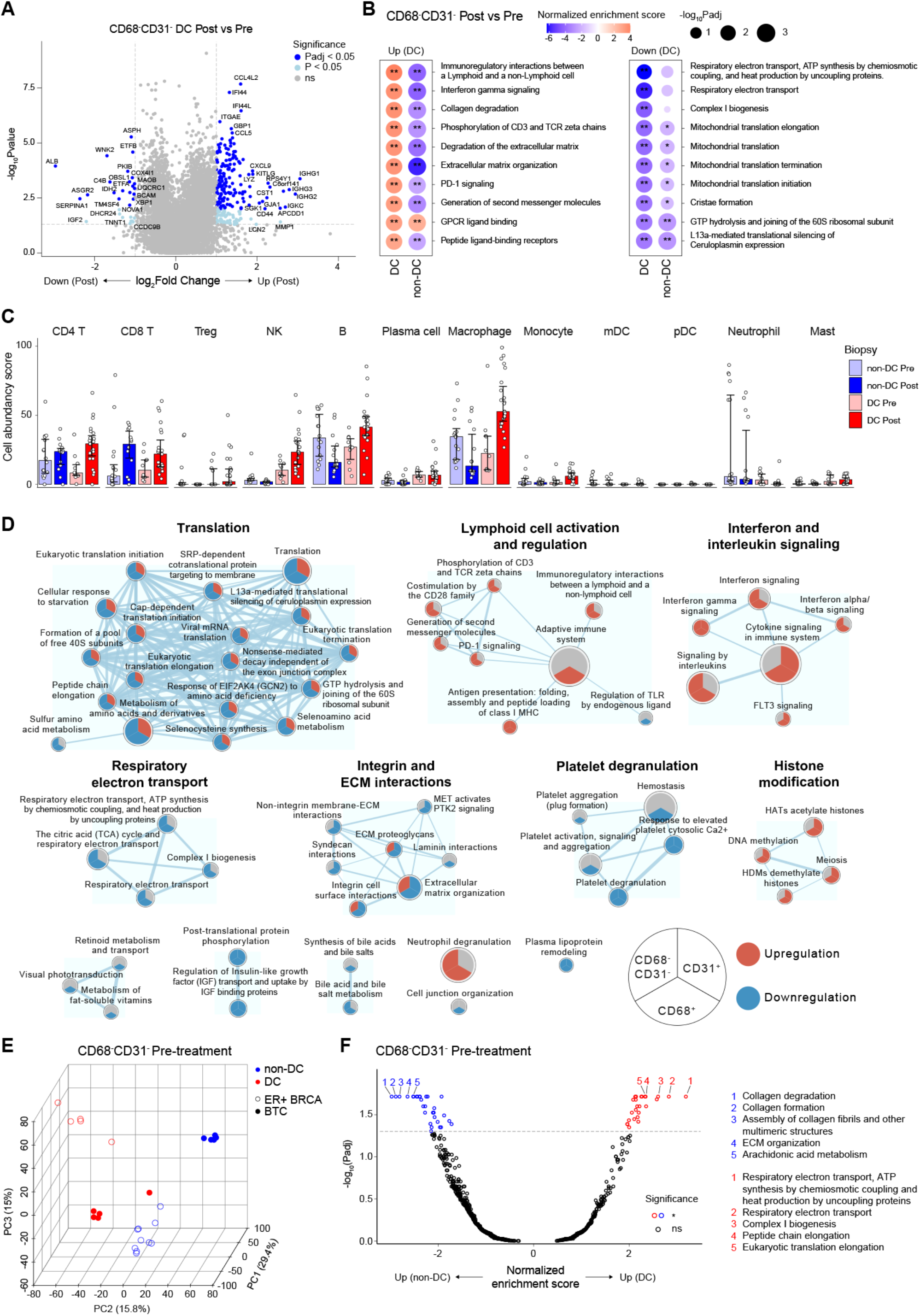
Tumor-infiltrating leukocyte recruitment and activation following bexmarilimab therapy in DC patients. **A**, Volcano plot showing differentially expressed genes in DC patient CD68^-^ CD31^-^ areas after bexmarilimab therapy. **B**, Bubble plots of top up- and downregulated pathways in CD68^-^CD31^-^ areas of DC patients after bexmarilimab therapy (gene set enrichment analysis). Red color denotes pathway activation and blue downregulation. **C**, Bar plots of cell abundancy scores calculated for the indicated immune cell types based on gene expression on CD68^-^CD31^-^ area. Median ± IQR, points represent individual ROIs. **D**, Enrichment map of significantly altered pathways (Padj < 0.05) in CD68^+^ areas of DC patients after bexmarilimab therapy. The map illustrates how these pathways have altered during bexmarilimab therapy on CD68^+^, CD31^+^ and CD68^-^CD31^-^ areas with red indicating significant upregulation, blue significant downregulation and gray non- significant change. Circle size represents pathway size and connecting line width represents the proportion of shared dataset genes between the two pathways. **E**, Principal component analysis of pre-treatment biopsy ROIs based on CD68^-^CD31^-^ area gene expression (n = 11,591 genes). **F**, Volcano plot showing differently activated pathways between CD68^-^CD31^-^ areas of non-DC and DC pre-treatment biopsies (gene set enrichment analysis). Top 5 pathways (lowest Padj) were annotated. BTC, biliary tract cancer; ER+ BRCA, estrogen receptor positive breast cancer; DC, disease control during bexmarilimab therapy; non-DC, no disease control during bexmarilimab therapy; NK, natural killer cell; mDC, myeloid dendritic cell; pDC, plasmacytoid dendritic cell; Treg, regulatory T cell; Pre, pre-treatment biopsy; Post, post-treatment biopsy; ROI, region of interest; **, Padj < 0.05; *, Padj < 0.05; ns, not significant.

The overall pathway changes in the analyzed segments visualized as an enrichment map showed that integrin and ECM interaction related pathways were downregulated specifically in the Clever-1^+^ tumor areas (CD68^+^ and CD31^+^), but not in the remaining CD68^-^CD31^-^ area, which reflects the aforementioned role of Clever-1 as an adhesion molecule and successful segmentation of these three areas (**Fig. 3D**). Furthermore, upregulation of interferon signaling and lymphocyte activation as well as downregulation of translation and respiratory electron transport were mutually observed in the CD68^+^ and CD68^-^CD31^-^ tumor areas, but not in the CD31^+^ vasculature (**Fig. 3D**).

To find common features of bexmarilimab responsive tumors, we compared pre-treatment biopsy gene expression between DC and non-DC patients. While the CD68^+^ biopsy areas were relatively similar between DC and non-DC patients (**Fig. 2H**), the CD68^-^CD31^-^ areas clustered based on the response rather than the cancer type and had a high number of differentially activated pathways between DC and non-DC patients (**Fig. 3E and F**). Higher expression of genes related to respiratory electron transport and lower expression of genes related to interleukin signaling in DC patient pre-treatment biopsies were observed (**Fig. 3F** and Supplementary Data Files). These observations suggest that the surrounding TME has a greater influence on bexmarilimab therapy outcome than existing TAM phenotypes. Collectively, these data illustrate that bexmarilimab therapy upregulates antigen presentation, T-cell activation and interferon signaling in tumors of DC patients, which results in enhanced chemokine production and immune cell recruitment into the TME.

## Discussion

Targeting macrophages to break the immune tolerance of tumors and help activate host immune defenses is the next cutting edge in cancer immunotherapy. Macrophage-targeted therapies could be used as monotherapy or along with various other treatments and open entirely new therapeutic options^16–18^. Here we report the first-in-human results of phase I/II MATINS study, evaluating bexmarilimab, a Clever-1 targeting antibody, in patients with advanced cancer, who have exhausted standard therapeutic options. Clever-1, a scavenger receptor, is highly expressed by immunosuppressive macrophages and targeting of the receptor with therapeutic antibodies, has previously been shown to result in re-programming of macrophages, reversion of macrophage mediated immunosuppression, revitalization of antigen presentation, and T-cell mediated anti-cancer immunity^8, 9, 11^. Thus, targeting Clever-1 represents a novel mechanism of action that has not been previously investigated as anti-cancer therapy.

Bexmarilimab administration was associated with favorable tolerability with no observed DLTs in part I, and MTD could not be defined. In part II, no additional safety signals were seen, and registered treatment-related AEs were typically low-grade. The observed excellent safety profile of the drug makes it feasible to test bexmarilimab in combination with other anti-cancer agents as well as to continue the development of bexmarilimab as a single-agent therapy. It is worth noting that several study patients developed immune-related AEs after relatively long exposure to bexmarilimab which were manageable with corticosteroids. Some of these patients also seemed to benefit from the treatment with partial response or prolonged tumor stabilization. Similarly, immune-related AEs have been linked to higher frequency of tumor responses in advanced cancer patients receiving PD-1 targeting antibodies^19^.

While efficacy was not the primary endpoint of the study, we found interesting preliminary evidence of anti-tumor activity for bexmarilimab. Although RECIST 1.1 defined ORR was low in the entire study population, promising DCR and long bexmarilimab treatment durations were observed in several patients, especially in cutaneous melanoma, gastric cancer, biliary tract cancer, HCC, and ER+ breast cancer. Treatment benefit measured by DC was also seen in anti-PD-(L)1 pretreated patients suggesting that single-agent bexmarilimab has activity also in immunotherapy refractory patients. No other clear association between the traditional baseline factors and DC were observed in the study. Tumor biomarker analysis showed an association between high baseline tumor Clever-1 expression and DC. Furthermore, circulating levels of TNFα (baseline) associated with DC and IFNγ increases were observed in DC patients. Low baseline circulating levels of IFNγ and tumor PD-L1 CPS score along with observed IFNγ increases in DC patients including those previously treated with immune checkpoint blockade inhibitors could indicate that bexmarilimab is able to rewire the macrophage mediated immunosuppression and result in T-cell mediated anti-tumor activity.

Indeed, when looking at macrophage activation signals by spatial transcriptomics there was a clear re-programming of TAMs in DC patients reflecting the findings observed in the peripheral biomarker analysis. The observed pro-inflammatory phenotype conversion, TFRC upregulation and downregulation of plasma lipoprotein remodeling was in line with the described changes in circulating monocytes after bexmarilimab therapy^11^, indicating that monocytes can retain at least some of the properties induced by bexmarilimab in circulation while differentiating into TAMs.

The immune activation was not restricted to TAMs as the neighboring cells showed robust signs of phosphorylation of CD3 and TCR zeta chains, and co-stimulation by the CD28 family suggesting that macrophages in these areas were in close contact with T-cells to facilitate their activation. It seemed that bexmarilimab increased immune cell abundance in tumors, which is contrary to published research describing impaired immune cell trafficking after Clever-1 blockade. While there is some selectivity in Clever-1 regulated trafficking^20^, a recent report by Steele and colleagues show that tumor lymphatics control the egress of T-cells after antigen exposure by ACKR3 upregulation, thus reducing CXCL12 sensitivity and promoting retention^21^. This might also be the case with Clever-1 targeting as we have previously shown that lymph node lymphatics in Clever-1 knock-out mice upregulate *Ackr2* after OVA-CFA administration^22^.

While the cell type deconvolution was performed for CD68^-^CD31^-^ area, some macrophage signal inevitably remained on this segment, facilitating macrophage abundancy estimation. Unexpectedly, we detected increased macrophage abundance after bexmarilimab therapy in DC patients. Inspection of representative CD68^+^ staining images confirmed this finding and we also observed upregulation of *CCL2* in CD68^+^ and CD68^-^CD31^-^ areas of DC biopsies. Typically, an increased number of TAMs would indicate poorer outcome. However, due to the conversion of macrophage phenotype with bexmarilimab the recruited monocytes were no longer immunosuppressive in nature and could support anti-tumor responses.

It still remains unclear what factors characterize patients who benefit from bexmarilimab therapy. The small spatial transcriptomics cohort did not allow us to make conclusive interpretations but gives insight into existing differences between the TME in DC and non-DC patients prior to therapy. In fact, it is surprising that even with this low number of samples significant differences can be observed between patients categorized as DC vs non-DC. The biomarker and profiling data, however, indicate that bexmarilimab would have better efficacy in tumors with low pre-existing interferon signatures. Since RECIST defined SD was observed with the study drug, additional analysis was carried out to characterize whether SD responses would be associated with signs of clinically meaningful, long-lasting anti-tumor activity. Interestingly, we observed a prolongation of survival in patients with PR/SD without this being associated with longer previous line of treatment duration, a surrogate for PFS. Furthermore, PFS2/PFS1 ratio of > 1.3 has often been proposed to indicate therapeutic benefit^23^ and this was observed significantly more frequently in PR/SD than PD patients. This is suggestive for the disease stabilizing effect of bexmarilimab, and even more importantly, a prolonged survival effect. A randomized trial is required to fully characterize the exploratory association between DC and survival.

The PK and PD profile of bexmarilimab was characterized by rapid elimination, receptor occupancy decay in circulating monocytes, and recovery of circulating soluble Clever-1 levels. Because of the faster than anticipated elimination of bexmarilimab, sampling carried out in part I did not enable us to define a comprehensive PK profile of the drug. We speculate that rapid elimination of bexmarilimab might be related to rapid internalization of bexmarilimab and its target as well as the short half-life of circulating monocytes (∼1d) and replenishment of new ones from the bone marrow. The short elimination of bexmarilimab did not, however, affect the immune activating pharmacodynamic effects. The further expansion cohorts of the MATINS trial will investigate more frequent dosing intervals and escalating dose levels to fully optimize the dosing and efficacy of bexmarilimab.

In conclusion, we present the results of part I and II of the MATINS trial, investigating a novel immunotherapeutic approach by targeting Clever-1 with the therapeutic antibody bexmarilimab. Bexmarilimab demonstrated an excellent safety and tolerability profile, and promising anti-tumor activity as monotherapy in several late-stage, treatment-refractory metastatic solid tumors. Further expansion of the MATINS study will investigate optimal dosing, biomarker-guided patient selection, and treatment efficacy, while separate novel trials will investigate bexmarilimab in earlier lines of treatment as well as in combination with other anti-cancer agents such as PD-(L)1 inhibitors.

## Methods

### Patients

This international, first-in-human, open-label, non-randomized phase I/II, dose escalation study was run according to Good Clinical Practice and the Declaration of Helsinki. The study was approved by local institutional review boards and national medicinal agencies (MATINS; NCT03733990, EudraCT 2018-002732-24). All participants provided written, informed consent before any trial-related investigations or treatment took place. Eligible patients had advanced (inoperable or metastatic), treatment-refractory, histologically confirmed hepatocellular carcinoma (HCC), gallbladder or intra- or extrahepatic biliary tract carcinomas (BTC), colorectal cancer (CRC), serous poorly differentiated ovarian cancer (OC), pancreatic ductal adenocarcinoma (PDAC), or immune checkpoint inhibitor refractory cutaneous melanoma without standard treatment options available. In addition, part II included patients with uveal melanoma, gastric adenocarcinoma, estrogen receptor positive (ER+) breast cancer, and anaplastic thyroid cancer (ATC). Patients were ≥ 18 years old with a life expectancy of ≥ 12 weeks, an Eastern Cooperative Oncology Group (ECOG) performance status of 0 or 1, adequate organ function, and no ongoing systemic infections, brain metastasis, history of autoimmune disease (except type I diabetes, celiac disease, hypothyroidism requiring only hormone replacement, vitiligo, psoriasis, or alopecia), use of concurrent antineoplastic therapies, or systemic steroids, and measurable disease (part II only).

### Procedures

The selection of the initial dose of 0.3 mg/kg was based on the nonclinical safety findings and affinity, binding, and receptor occupancy of bexmarilimab to Clever-1 and monocytes *in vitro*. No relevant toxicity was observed with 100 mg/kg dosing in non-human primates and 0.3 mg/kg dose should result in Clever-1 receptor occupancy in monocytes. Dose escalation for 0.1, 0.3, 1, 3, and 10 mg/kg (every 3 weeks (Q3W) intravenously) were characterized by two-stage time-to-event continual reassessment method (TITE-CRM) which allowed for more accurate determination of the MTD and staggered patient accrual without the need for complete DLT follow-up of previously treated patients, compared to conventional dose-finding designs^24^. Adverse events (AE) were collected according to the National Cancer Institute-Common Terminology Criteria for Adverse Events (NCI-CTCAE) version 5.0.

### Outcomes

The primary objectives of part I of the study were to characterize safety, tolerability, MTD, and define the recommended dose of bexmarilimab for parts II and III of the trial. Secondary objectives included pharmacokinetic (PK) profiling, evaluation of immunogenicity of bexmarilimab, and preliminary efficacy according to the objective response rate (ORR) and immune-related ORR (irORR). DLTs were defined as a treatment-related grade ≥ 3 AE or laboratory abnormality occurring ≤ 63 days following the first dose of bexmarilimab with exceptions of grade 3 infusion reactions, nausea/vomiting, thrombocytopenia, or neutropenia. Furthermore, grade 2 AST/ALT accompanied with 2x ULN elevation of bilirubin (with normal AST/ALT at baseline), or dose delay of second cycle of bexmarilimab due to drug-related toxicity by ≥ 14 days were considered as DLTs. In part II, the primary objectives were safety, tolerability and preliminary efficacy of bexmarilimab monotherapy with ORR, DCR, and irORR in distinct cancer type specific expansion groups. Secondary objectives included Clever-1 expression in each tumor type, PK profile and immunogenicity of bexmarilimab, predictive biomarkers of efficacy, and duration of response.

### Pharmacokinetics, Receptor Occupancy and Tumor Biomarker analysis

The PK profile of a single dose (during Cycle 1) and repeated doses (during Cycles 1-5) of bexmarilimab were determined by repeated measurements of the drug concentration in the circulation. Peak concentration (Cmax), through concentration (Cmin), area under the plasma concentration versus time curve (AUC), clearance, volume of distribution, terminal half-life (t1/2), receptor occupancy (RO) on circulating monocytes and circulating soluble Clever-1 levels at each dose level were determined. Pre-treatment tumor samples were immunohistochemically (IHC) stained for Clever-1 (4G9) and PD-L1 (22C3). Clever-1 was scored as the percentage (0-100%) of positive viable cells (irrespective of location, intra-tumor, or stroma) while PD-L1 with combined positivity score (CPS). Circulating cytokines were analyzed using a panel approach. All methods on PK, RO, and biomarker analysis are provided in supplemental appendix.

### GeoMx Digital Spatial Profiling of MATINS biopsies

The spatial gene expression profiles of MATINS pre- and post-treatment biopsies were analyzed using GeoMx Digital Spatial Profiler (NanoString) with NGS readout. The method uses immunofluorescence staining to annotate tissue morphology and RNA probes coupled to photocleavable oligonucleotide tags to measure gene expression. Oligonucleotide tags are released from selected tissue regions using UV exposure (area of illumination, AOI) and quantitated by next-generation sequencing. Tissue sections (5 µm) from tumor samples were mounted on Superfrost slides and baked at 60°C in a drying oven for 1 hour. After deparaffinization and rehydration, the protein targets were retrieved by dipping the slides into DEPC-treated water at 100°C for 10 seconds, followed by incubating the slides in 1X Tris-EDTA (pH 9.0) at 100°C for 20 minutes without further heating. The slides were then washed with phosphate-buffered saline (PBS) at room temperature for 5 minutes. To expose the RNA targets, the slides were next incubated in preheated Proteinase K solution (0.1 µg/mL) at 37°C for 20 minutes and washed with PBS for 5 minutes, followed by incubation in 10% neutral buffered formalin (NBF) for 5 minutes, NBF-stop buffer (2 x 5 minutes), and washing with PBS for 5 minutes, all at room temperature.

Hybridization was performed using the GeoMx Human Whole Transcriptome Atlas (cat. GMX-RNA-NGS-HuWTA) kit following the manufacturer’s instructions. Briefly, Atlas probe mix was diluted in buffer R (pre-heated to 37°C) and applied to the slides. The sections were then covered with a Grace Bio-Labs HybriSlip for the hybridization reaction and incubated at 37°C overnight (16 hours) in a hybridization oven. The following day, the slides were washed with 2X saline-sodium citrate (SSC), 2XSSC-T (20xSSC+0.7ml 10% Tween-20+62.3ml DEPC-treated water), and 2 x 2XSSC-50% formamide to remove the HybriSlip and reduce the binding of off-target probes, and moved back into 2XSSC buffer.

For the morphology staining, sections were blocked with Buffer W at room temperature for 30 minutes (protected from light) and stained with SYTO 83 (Invitrogen, cat #: S11364, 1:35000), PanCK (Novus Biologicals, cat #: NBP2-33200af488, 1:200), CD68 (Santa Cruz, cat #: sc- 20060af594, 1:40), and CD31 (Abcam, cat #: ab215912, 1:110) morphology markers diluted in buffer W for 2 hours in a humid chamber at room temperature (protected from light). After staining, the sections were washed with 2X SSC for 2 times and immediately loaded on the GeoMx DSP.

The ROIs were selected based on the abundance of CD68 and CD31 staining in pan-cytokeratin rich areas to achieve at least 50 cells in each segment. The segmentation of ROIs into the three areas of illumination (AOI) was performed in following order: 1.) selection of CD68^+^pan-cytokeratin^-^ area, 2.) selection of CD31^+^ area and 3.) selection of the remaining DNA^+^ area. These three non-overlapping areas are referred as CD68^+^, CD31^+^ and CD68^-^CD31^-^ throughout the text.

Image scan and probe tag collection were performed using the GeoMx Digital Spatial Profiler (NanoString) instrument. The resulting libraries were sequenced on Illumina NovaSeq 6000 system with following read length configuration: R1: 27, i7: 8, i5: 8 and R2: 27. The sequencing data was demultiplexed and converted into FASTQ files with bcl2fastq2 Conversion Software (Illumina, v2.2.0). Raw FASTQ files were subjected to adapter removal, merging of paired-end reads, read alignment to barcode IDs and PCR duplicate removal with GeoMx NGS Pipeline software (NanoString), and resulting DCC files were uploaded into GeoMx DSP Control Center (Nanostring, v.2.4). High-resolution .tif images of ROIs were exported from GeoMx DSP Control Center to ImageJ (NIH, USA, v1.53q) and ROIs with representative quantity of CD68^+^ cells were displayed by selecting 400µm × 400µm regions (limited by smallest ROI) from the ROI center.

### GeoMx NGS readout data analysis

GeoMx analysis workflow consisting of QC, normalization, differential gene expression analysis and pathway analysis was run in GeoMx DSP Control Center according to NanoString’s recommendations (GeoMx-NGS Data Analysis User Manual, SEV_00090-05 for software v2.4). Subsequent analyses and visualizations were performed with R (v.4.0.4, tidyverse v.1.3.1)^25, 26^. For QC, negative probes identified as outliers by Grubbs test were excluded in each segment. Signal-to-noise ratio [Q3 / geoMean(Negative probes)] was calculated for each segment and segments with signal-to-noise ratio ≤ 1 were excluded from further analyses. Genes exceeding both the limit of quantitation calculated based on negative probe counts and value 2 in at least 15% of segments were defined as confidently expressed and used in further analyses. For analyses not directly comparing different segment types (differential gene expression and pathway analyses), this gene filtering was performed separately for CD68^+^, CD31^+^ and CD68^-^CD31^-^ segments. Filtered data were normalized by Q3 values calculated based on all genes passing the aforementioned filters, and normalized counts used in all further analyses and visualizations.

Unsupervised hierarchical clustering and principal component analyses were performed with R (functions hclust and prcomp, respectively) using log2-transformed normalized counts. Hierarchical clustering was visualized as a heatmap using ComplexHeatmap package (v2.6.2)^27^ and first three principal components as a 3D scatterplot using scatterplot3d package (v0.3-41)^28^. Differentially expressed genes (DEGs) were identified using linear mixed models to account for multiple observations (ROIs) from the same biopsy. DEGs between post- and pre-biopsies were identified separately for DC and non-DC group using a linear mixed model with biopsy type as a fixed effect and patient ID as a random effect. DEGs between DC and non-DC pre-biopsies were identified similarly by using a linear mixed model with response group as a fixed effect and patient ID as a random effect. Obtained p-values and log2Fold Changes were plotted as volcano plots and genes passing FDR threshold 0.05 (Benjamini-Hochberg) were indicated by point color. Significantly altered pathways between the indicated conditions were identified with GeoMx DSP Control Center that performs gene set enrichment analyses using fGSEA package^29^ and Reactome pathway database (v78). Pathways with <20% coverage (% of pathway genes expressed in the dataset) were excluded from the analysis, resulting in 73.2% median coverage. Obtained normalized enrichment scores and adjusted p-values were plotted as volcano plots or as bubble plots with dot area corresponding to - log10Padjusted value. Pathways with adjusted P value lower than 0.05 were visualized as an enrichment map using Cytoscape’s (v3.9.1)^30^ Enrichment Map app (v3.3.4)^31^ with yFiles Organic Layout. The created network was clustered by applying Markov clustering algorithm with default settings and resulting clusters were manually annotated based on annotations created with Cytoscape’s AutoAnnotate app (v1.4.0)^32^.

To evaluate TAM phenotypes, M1 and M2 macrophage scores were calculated for each CD68^+^ segment based on genes differentially expressed in M1 and M2 macrophages^13^ (Table 1 in Martinez et al.) and passing our aforementioned gene filtering criteria. The expression levels of these M1 and M2 genes were plotted as a heatmap after log2-transformation and z-scoring. M1 and M2 scores were calculated separately for each CD68^+^ segment by dividing average M1 (or M2) gene expression level by average overall gene expression level. For evaluating the abundancies of different immune cell types within the CD68^-^CD31^-^ biopsy areas, we used SpatialDecon package (v1.0.0)^33^ and its safeTME tumor-immune deconvolution cell profile matrix with default settings, except for providing safeTME cell type matches for major cell type calculation and raw count matrix for data-point weighting.

### Statistical analysis

The analysis populations are described in detail in the supplementary appendix. The data cut-off date for analysis was Jan 31^st^, 2022. ORR and DCR were evaluated at cycle 4 according to RECIST 1.1. PFS was evaluated from the date of the first dose of bexmarilimab until documented disease progression, death, or end of follow-up, the former two counted as events. OS was calculated from the date of the first dose of bexmarilimab until death or end of follow-up, the former was counted as an event. Previous line of therapy duration was calculated from the first treatment date until therapy discontinuation. Landmark analysis for OS was analyzed from the fourth study treatment cycle date until death or end of follow-up, the former was counted as an event. PFS and OS were analyzed using the Kaplan-Meier method and Cox-regression with 95% confidence interval. For the IHC biomarker analysis, Wilcoxon two-sample test was applied. For the PFS2/PFS1 ratio, percentage of patients with ratio of > 1.3 were scored positive^23^. The data were analyzed by a statistician employed by the sponsor and by the senior academic authors.

## Data Availability

All data produced in the present study are available upon reasonable request to the authors

## Availability of data and material

Access to participant-level data is currently not available.

## Competing interests

LV declares consulting fees from Adaptherapy; Stock or stock options for Resolved.

MdM declares payment or honoraria for lectures, presentations, speakers bureaus, manuscript writing or educational events from Janssen and MSD

AP declares payment or honoraria for lectures, presentations, speakers bureaus, manuscript writing or educational events from Incyte and Roche; Support for attending meetings and/or travel from Gilead; Participation on a Data Safety Monitoring Board or Advisory Board for Incyte, Novartis, Gilead, and BeiGen.

DB declares consulting fees from Merck AG, Pfizer, Bayer, Cantargia AB, Faron Pharmaceuticals, and Servier.

TS declares payment or honoraria for lectures, presentations, speakers bureaus, manuscript writing or educational events from BMS; Participation on a Data Safety Monitoring Board or Advisory Board for Faron Pharmaceuticals, Merck, and Novartis; Leadership or fiduciary role in other board, society, committee or advocacy group, paid or unpaid for Finnish Oncology Society.

SI declares grants or contracts from any entity from Roche and AstraZeneca; Payment or honoraria for lectures, presentations, speakers bureaus, manuscript writing or educational events from BMS, Roche, Astra Zeneca, Takeda, Novartis, MSD, Pierre Fabre, Boehringer-Ingelheim.

SS declares consulting fees from Faron Pharmaceuticals, Participation on a Data Safety Monitoring Board or Advisory Board for Faron Pharmaceuticals.

YTM declares consulting fees from Eisai, Roche, AstraZeneca, Ipsen; Payment or honoraria for lectures, presentations, speakers bureaus, manuscript writing or educational events for Bayer, Boston Scientific; Participation on a Data Safety Monitoring Board or Advisory Board for Faron Pharmaceuticals.

DG declares consulting fees from Clinigen and McCann Health; Payment or honoraria for lectures, presentations, speakers bureaus, manuscript writing or educational events from Cancer Drug Development Fund and Pfizer.

SPA declares payment or honoraria for lectures, presentations, speakers bureaus, manuscript writing or educational events from Bayer, BMS, ASCO, and Exelixis; Participation on a Data Safety Monitoring Board or Advisory Board for AstraZeneca, Seagen, and QED Therapeutics/Helsinn; Leadership or fiduciary role in other board, society, committee or advocacy group, paid or unpaid for Board-Support New India.

PJ declares Participation on a Data Safety Monitoring Board or Advisory Board for Faron Pharmaceuticals.

CY declares consulting fees from Faron Pharmaceuticals; Payment or honoraria for lectures, presentations, speakers bureaus, manuscript writing or educational events from Bayer.

JM declares all support for the present manuscript from Faron Pharmaceuticals (employment); Patents planned, issued or pending for Faron Pharmaceuticals; Participation on a Data Safety Monitoring Board or Advisory Board for Faron Pharmaceuticals; Stock or stock options for Faron Pharmaceuticals.

MKK declares all support for the present manuscript from Faron Pharmaceuticals (employment); Patents planned, issued or pending for Faron Pharmaceuticals; Stock or stock options for Faron Pharmaceuticals.

JJ declares all support for the present manuscript from Faron Pharmaceuticals (employment); Grants or contracts from any entity from European Innovation Council; Stock or stock options for Faron Pharmaceuticals.

JPK declares all support for the present manuscript from Faron Pharmaceuticals; Consulting fees from MSD, BMS, Roche, Pfizer, and AstraZeneca; Payment or honoraria for lectures, presentations, speakers bureaus, manuscript writing or educational events from MSD, BMS, Roche, AstraZeneca, and Sanofi; Payment for expert testimony from Sanofi; Support for attending meetings and/or travel from BMS; Participation on a Data Safety Monitoring Board or Advisory Board for Faron Pharmaceuticals; Stock or stock options from Faron Pharmaceuticals.

AM declares payment or honoraria for lectures, presentations, speakers bureaus, manuscript writing or educational events from Janssen, Chugai, Novartis, and Bayer; Support for attending meetings and/or travel from Amgen; Participation on a Data Safety Monitoring Board or Advisory Board for Janssen, Takeda, Genmab, Merck, and Faron Pharmaceuticals.

MH declares all support for the present manuscript from Faron Pharmaceuticals; Consulting fees from Faron Pharmaceuticals; Stock or stock options from Faron Pharmaceuticals PB declares consulting fees from Faron Pharmaceuticals, Herantis Pharma, MSD Oncology, Ipsen, Oncorena, TILT biotherapeutics; Participation on a Data Safety Monitoring Board or Advisory Board for Faron Pharmaceuticals, TILT biotherapeutics and Oncorena; Leadership or fiduciary role in other board, society, committee or advocacy group, paid or unpaid for Terveystalo (employment); Stock or stock options for Terveystalo, TILT biotherapeutics; Other financial or non-financial interests for Faron pharmaceuticals, stock ownership (spouse)

## Funding

This study was funded by Faron Pharmaceuticals (Turku, Finland), European Union’s Horizon 2020 research and innovation program (ID: 960914), the Academy of Finland (MH), Business Finland (MH), Cancer Foundations (MH and JHR), the Sigrid Jusélius foundation (MH), Orion Research Foundation (JHR) and the Paulo Foundation (JHR).

## Authors’ contributions

JHR, SS, CY, JM, MKK, JJ, JPK, MH and PB designed the study. LV, MDM, AP, DR, TS, SI, YTM, DG, SPA, PJ, AM recruited and treated the patients, and collected the data. JHR, YX, SK and MH optimized and performed GeoMx. JHR, CY, JM, MKK, JJ, JPK, MH, and PB analysed and interpreted the data. JHR, JPK, MH and PB wrote the first draft of the manuscript. All authors reviewed the manuscript and approved the final version.

## Acknowledgements

We thank the study participants, the investigators, and the research team who contributed to the study. We also want to thank Mari Parsama, Teija Kanasuo, Riikka Sjöroos, Sari Mäki and Maritta Pohjansalo for excellent technical assistance. GeoMx Digital Spatial Profiling was performed at FIMM Single-Cell Analytics and Sequencing units supported by HiLIFE and Biocenter Finland.

## Notes

### Clinical Trial

NCT03733990

### Funding Statement

This study was funded by Faron Pharmaceuticals (Turku, Finland), European Unions Horizon 2020 research and innovation program (ID: 960914), the Academy of Finland, Business Finland, Cancer Foundations, the Sigrid Juselius foundation, Orion Research Foundation and the Paulo Foundation.

### Author Declarations

Local institutional ethical committees of Oulu University Hospital, Helsinki University Hospital, Tampere University Hospital, Turku University Hospital, University Hospitals Birmingham, Institut Gustave Roussy, Royal Marsden Hospital, Erasmus Medical Center, The Christie NHS Foundation Trust and Mays Cancer Center gave ethical approval for this work. The study was approved by national medicinal agencies (MATINS; NCT03733990, EudraCT 2018-002732-24).

