## Supplementary material for "Bexmarilimab-induced macrophage activation leads to treatment benefit in solid tumors: the phase I/II first-in-human MATINS trial": Supplemetary material

### Supplementary appendix

#### List of MATINS investigators

Finland: Petri Bono, Annika Pasanen, Tanja Skyttä, Sanna Iivanainen, Panu Jaakkola
France: Loic Verlingue
Netherland: Debbie Robbrecht, Maja De Jonge
Spain: Maria de Miguel
UK: Anna Minchom, Shishir Shetty, Yuk Ting Ma, Donna Graham
USA: Sukeshi Arora

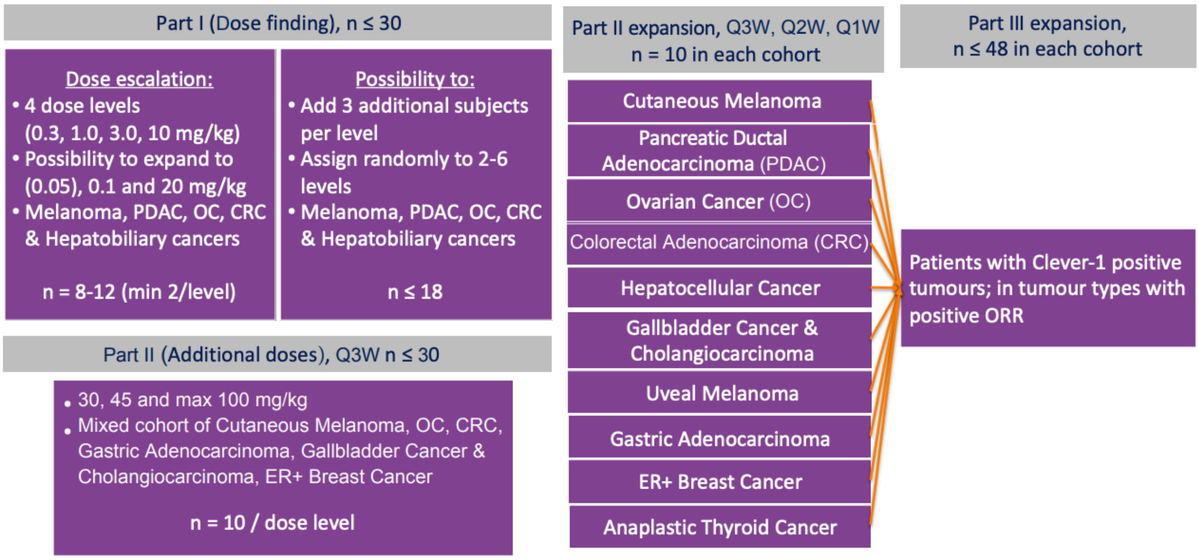

Figure S1: MATINS study design

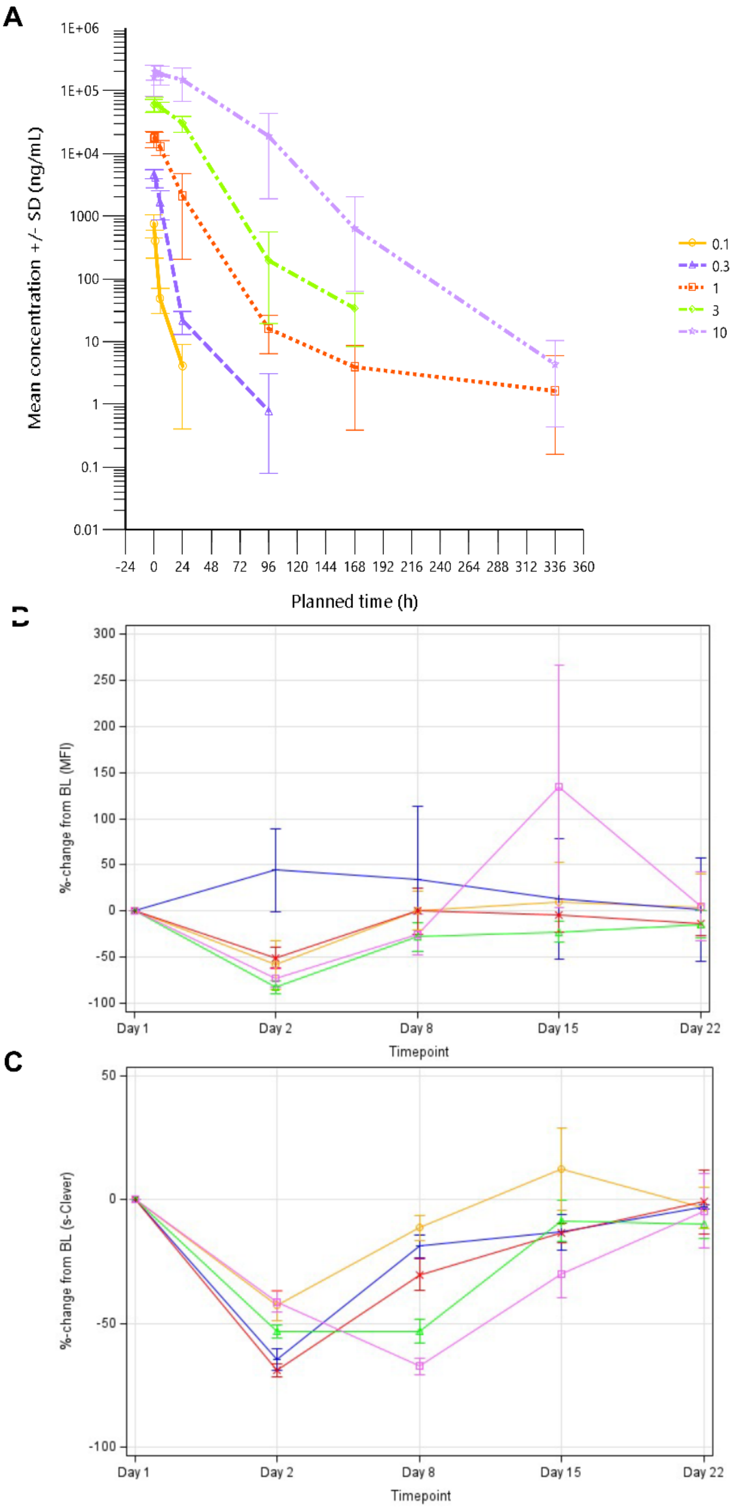

Figure S2: Pharmacokinetics, receptor occupancy, and soluble clever-1 after the first dose bexmarilimab

(A) Pharmacokinetics of bexmarilimab. X-axis is showing the concentration of bexmarilimab while Y-axis presents the time in hours from the IMP infusion. (B) Receptor occupancy for Clever-1 on circulating monocytes. X-axis is percentile change from baseline while Y-axis the time in days from the IMP infusion. (C) Soluble Clever-1 level. X-axis is percentile change from baseline while Y-axis the time in days from the IMP infusion. Investigated dose levels were 0.1 (yellow), 0.3 (blue), 1 (red), 3 (green), and 10mg/kg (purple). BL=Baseline, s-Clever = soluble clever-1. MFI=Mean fluorescence index.

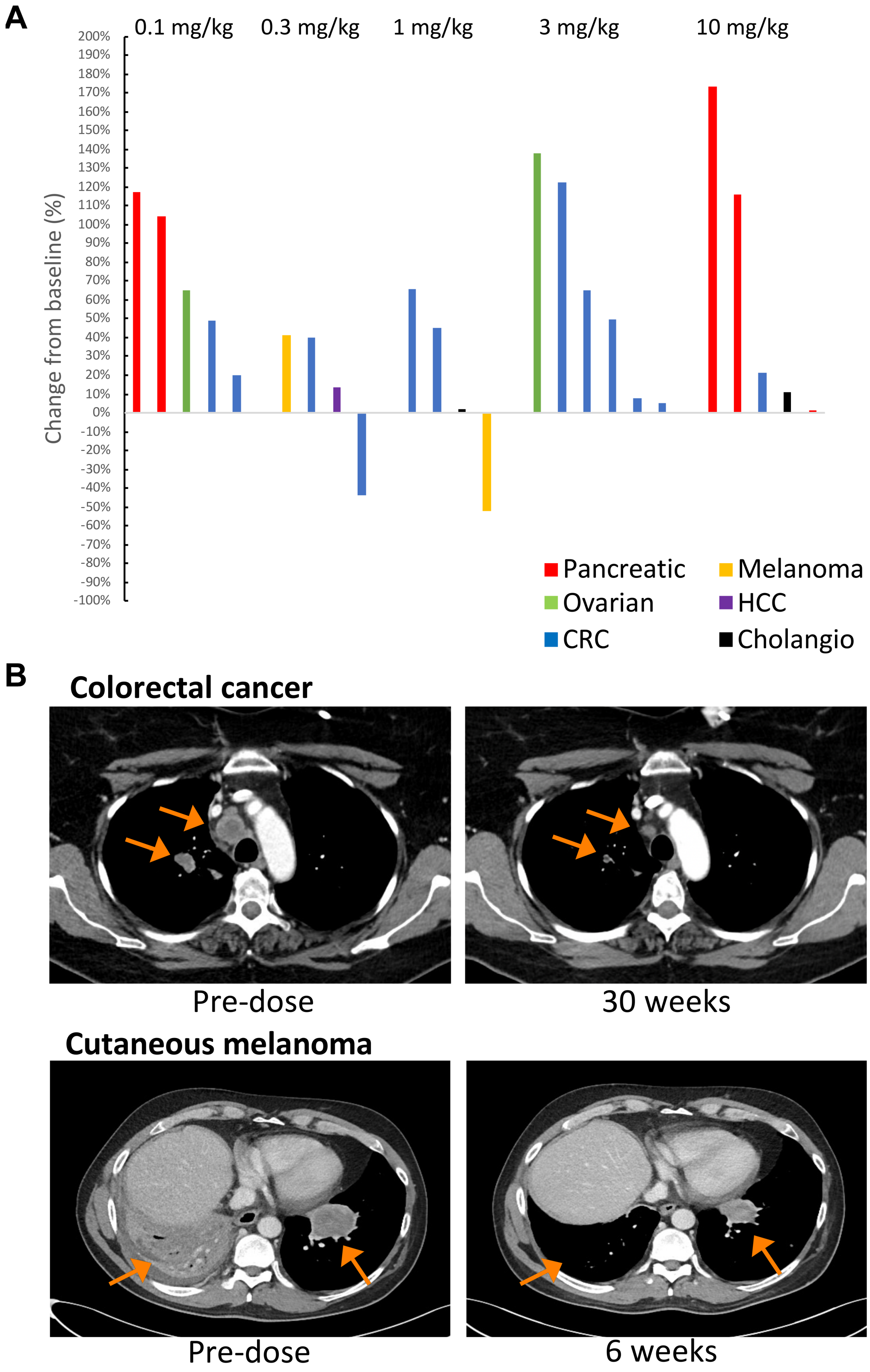

Figure S3: Preliminary anti-tumor efficiency for bexmarilimab at part I

(A) Waterfall plot for the best target lesion responses (%) according to dose and tumor type in RECIST 1.1 evaluable patients (n=23). (B) CT-scans for the selected responding patients in baseline and at the time of the best response. Colorectal cancer patient with PR response and melanoma patient with PR-response in target lesions are presented.

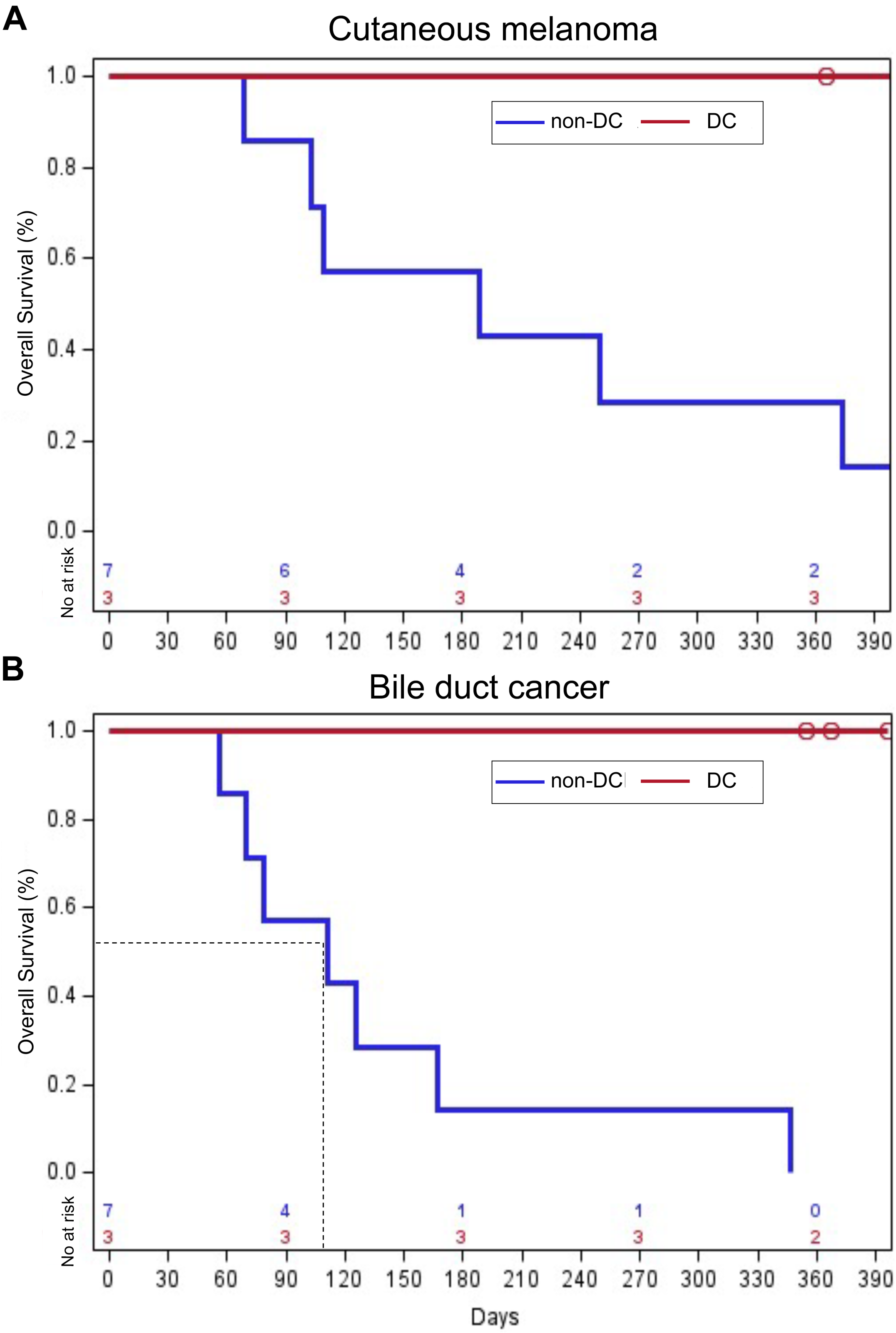

Figure S4: Overall survival analysis according to DC in cutaneous melanoma and bile duct cancers

Circles indicate sensored events.

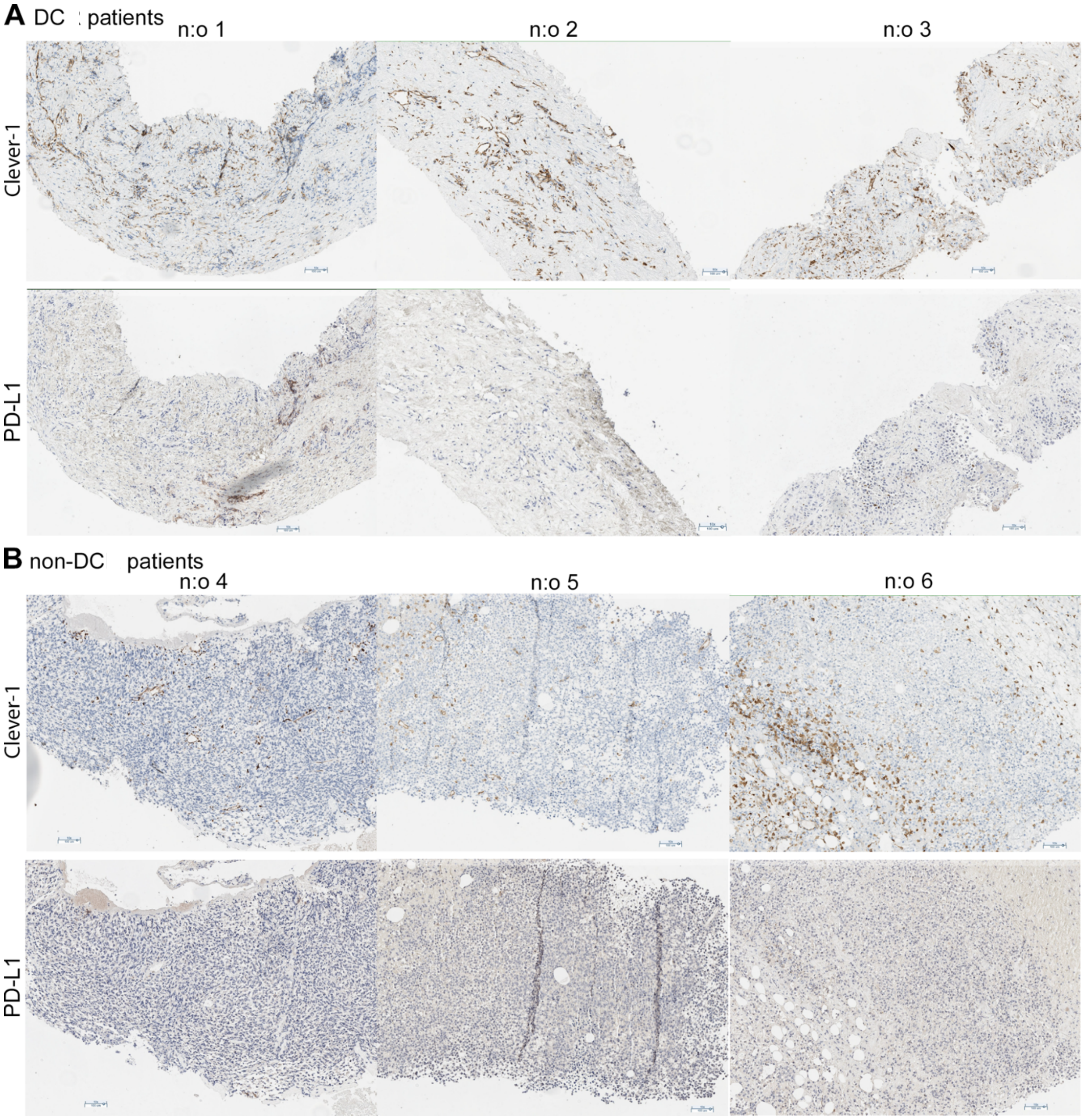

Figure S5: Tumor Clever-1 and PD-L1 expression in selected DC and non-DC patients

Immunohistochemical staining of pre-treatment tumor samples (A) Clever-1 and PD-L1 for selected DC patients (1-3) (B) Clever-1 and PD-L1 for selected non-DC subject (4-6). Line segment for 100μm.

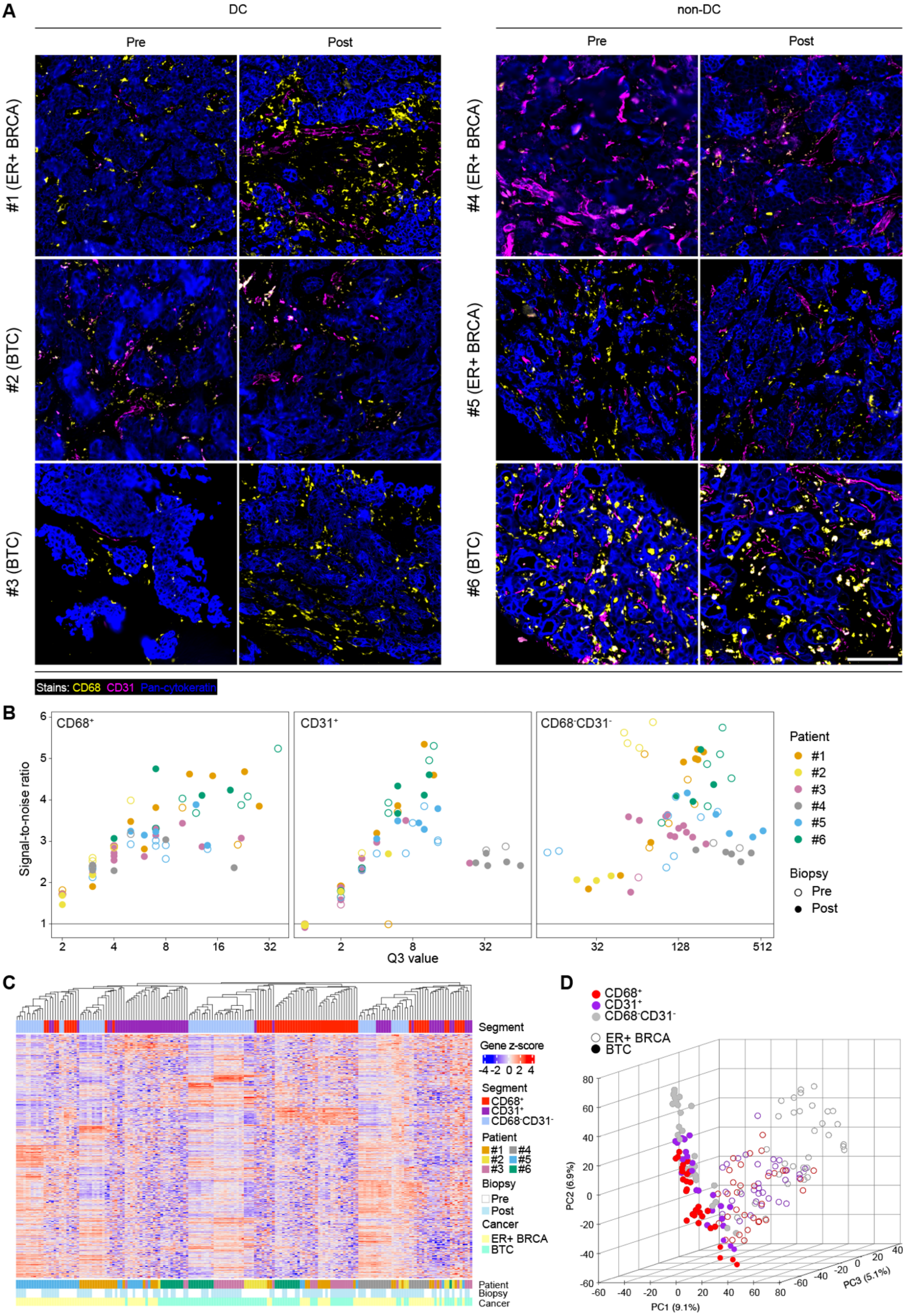
A
DC
non-DC
Pre
Post
Pre
Post
#1 (ER+ BRCA)
#2 (BTC)
#3 (BTC)
#4 (ER+ BRCA)
#5 (ER+ BRCA)
#6 (BTC)
Stains: CD68 CD31 Pan-cytokeratin
B
CD68+
CD31+
CD68-CD31-
Signal-to-noise ratio
Q3 value
Patient
#1
#2
#3
#4
#5
#6
Biopsy
Pre
Post
C
Segment
Gene z-score
-4 -2 0 2 4
Segment
CD68+
CD31+
CD68-CD31-
Patient
#1 #4
#2 #5
#3 #6
Biopsy
Pre
Post
Cancer
ER+ BRCA
BTC
Patient
Biopsy
Cancer
D
CD68+
CD31+
CD68-CD31-
ER+ BRCA
BTC
PC2 (6.9%)
PC1 (9.1%)
PC3 (5.1%)

Figure S6: GeoMx spatial transcriptomics profiling of pre- and post-treatment biopsies from MATINS trial patients.

**A**, Morphology marker staining for GeoMx analysis showing $CD68^{+}$ macrophages (yellow), $CD31^{+}$ vessels (magenta) and pan-cytokeratin$^{+}$ cancer cells (blue). Images of representative ROIs were selected based on CD68 staining in each biopsy and 400µm × 400µm square regions are displayed from the center of the ROIs. Scale bar 100 µm. **B**, Signal-to-noise ratio (Q3 value / geoMean[NegativeProbes]) is shown separately for $CD68^{+}$, $CD31^{+}$ and $CD68^{-}CD31^{-}$ segments with points representing analyzed ROIs. Segments with signal-to-noise ratio ≤1 were excluded from further analyses. **C** and **D**, Clustering of all QC-passing segments based on Q3-normalized and $\log_2$-transformed counts (n = 10,612 genes). Heatmap of unsupervised hierarchical clustering with columns representing segments (**C**) and a scatter plot of the first three principal components with each point representing a single segment (**D**). BTC, biliary tract cancer; ER+ BRCA, estrogen receptor positive breast cancer; DC, disease control during bexmarilimab therapy; non-DC, no disease control during bexmarilimab therapy; Pre, pre-treatment biopsy; Post, post-treatment biopsy; Q3, $75^{th}$ percentile of counts; ROI, region of interest.

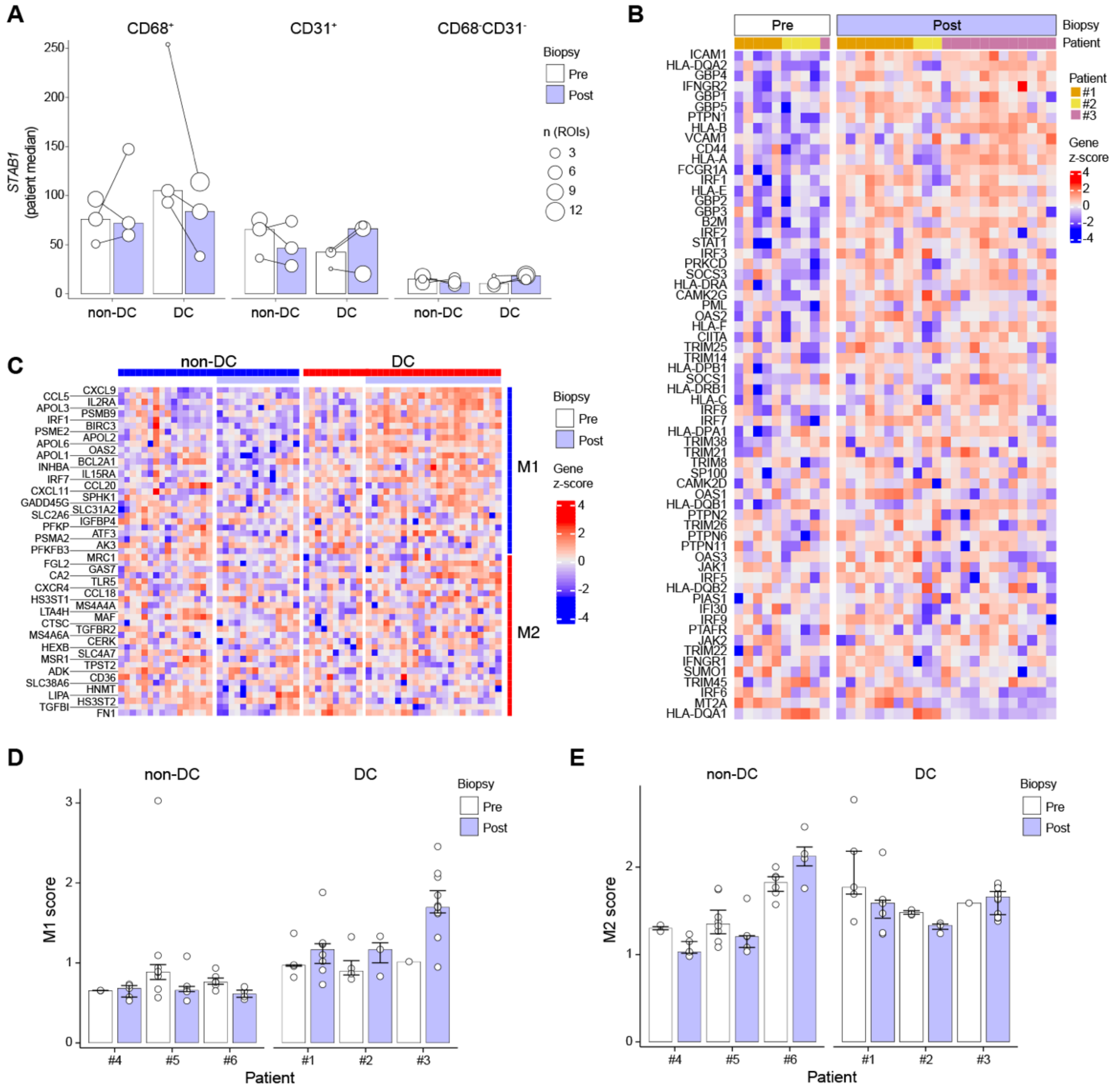

Figure S7: GeoMx profiling of biopsy CD68+ area transcriptome after bexmarilimab therapy.

**A**, Clever-1 mRNA (*STAB1*) levels in CD68$^+$, CD31$^+$ and CD68$^-$CD31$^-$ areas of DC and non-DC patient biopsies. Points indicate median expression across patient's ROIs and bars represent patient group median. **B**, Expression levels of interferon gamma signaling pathway genes measured from CD68$^+$ areas of DC patient biopsies. **C**, Heatmap of M1 and M2 macrophage marker gene expression levels calculated from CD68$^+$ biopsy areas. **D** and **E**, Bar graphs of M1 (**D**) and M2 (**E**) scores calculated based on mean expression level of macrophage marker genes shown in **C**. Score of 1 indicates marker gene expression level equal to overall gene expression level on CD68$^+$ biopsy area. Median ± IQR, points represent individual ROIs. DC, disease control during bexmarilimab therapy; non-DC, no disease control during bexmarilimab therapy; Pre, pre-treatment biopsy; Post, post-treatment biopsy; ROI, region of interest. In **B** and **C**, color gradient represents gene z-scores calculated from Q3-normalized and $log_2$-transformed counts with red corresponding to higher expression.

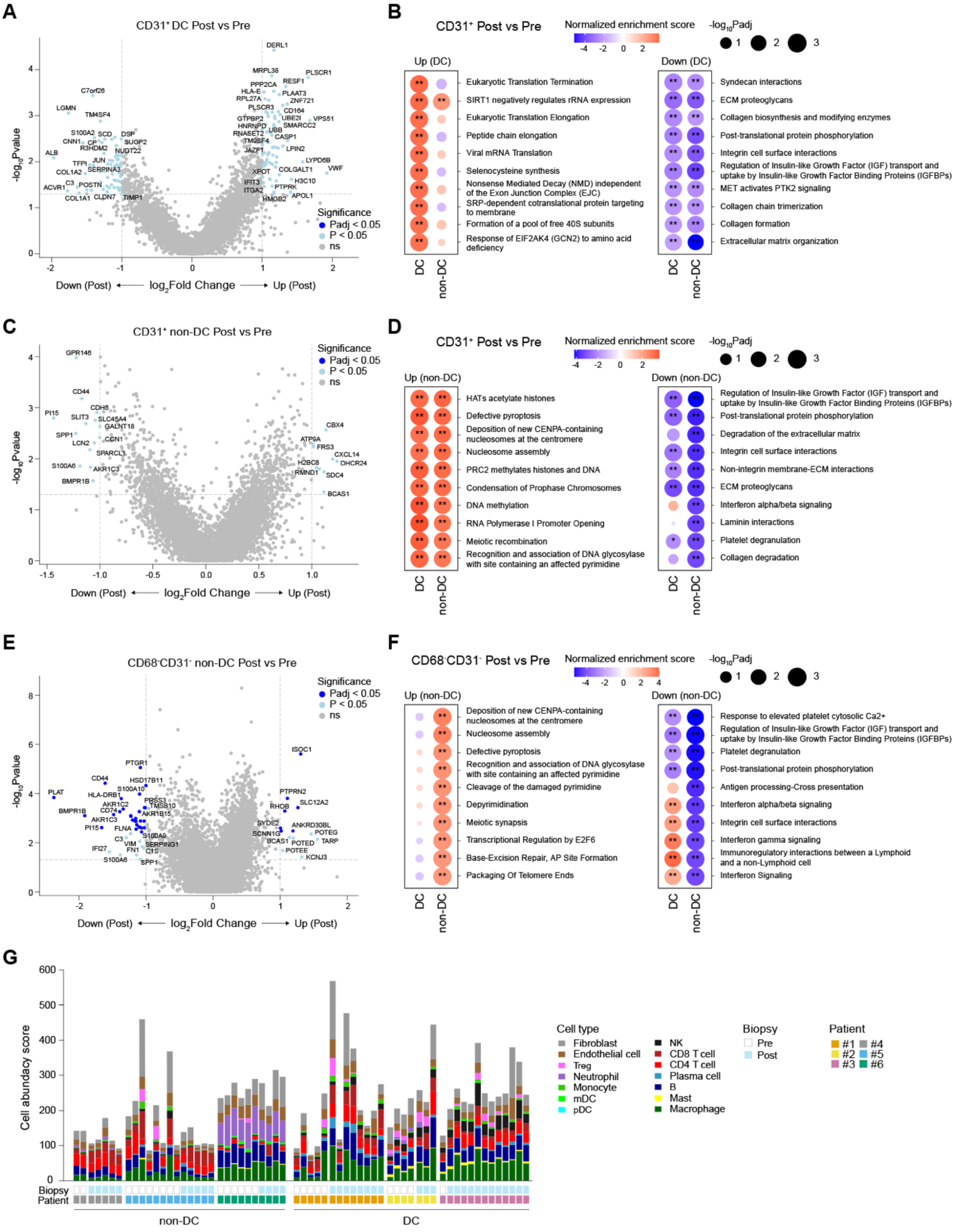

Figure S8. GeoMx profiling of CD31+ and CD68-CD31- tumor area transcriptomes after bexmarilimab therapy. **A** to **F**, Analysis of gene expression changes after bexmarilimab therapy in CD31$^+$ tumor areas of DC patients (**A** and **B**), CD31$^+$ tumor areas of non-DC patients (**C** and **D**) and CD68$^-$CD31$^-$ tumor areas of non-DC patients (**E** and **F**). Volcano plots show differentially expressed genes (**A**, **C** and **E**) and bubble plots top up- and downregulated pathways (**B**, **D** and **F**; gene set enrichment analysis). In **B**, **D**, and **F**, red color denotes pathway activation and blue downregulation. **G**, Cell abundancy scores for the indicated cell types were calculated based on Q3-normalized gene expression on CD68$^-$

CD31$^{-}$ area using SpatialDecon. Each stacked bar represents a single ROI. DC, disease control during bexmarilimab therapy; non-DC, no disease control during bexmarilimab therapy; Pre, pre-treatment biopsy; Post, post-treatment biopsy; ROI, region of interest. *, FDR < 0.05; **, FDR < 0.01; ns, not significant.

#### Table S1: Inclusion and exclusion criteria

**Inclusion Criteria**

1. Written Informed Consent
2. Aged ≥ 18 years male or female
3. Tumour sample should be collected during screening period. If a recent tumour biopsy obtained within six months before the date of consent is available (or older, as agreed on a case by case basis with the sponsor), that may be used. At the discretion of the sponsor, the tumour sample may be optional for certain subjects in Part III
4. Life expectancy > 12 weeks
5. Histologically confirmed advanced (inoperable or metastatic) malignancies in which (according to the view of the investigator) no curative, effective or suitable treatment options exist.
   - Hepatocellular carcinoma
   - Gallbladder cancer or intra- or extrahepatic cholangiocarcinoma
   - Colorectal adenocarcinoma
   - Serous poorly differentiated (Grade 3) ovarian adenocarcinoma or undifferentiated ovarian cancer
   - Pancreatic ductal adenocarcinoma
   - Immunotherapy (IO) resistant cutaneous melanoma (progression during programmed cell death protein-1 (PD-1)/programmed cell death ligand-1 (PD-L1) or CTLA-4 antibody therapy)
   - Uveal melanoma in Parts II and III
   - Gastric adenocarcinoma (including adenocarcinoma of the distal esophagus / GE junction) in Parts II and III
   - ER+ breast cancer in Parts II and III
   - Anaplastic thyroid cancer in Parts II and III
6. Eastern Cooperative Oncology Group (ECOG) performance status 0 or 1
7. Measurable disease in Parts II and III
8. Adequate bone marrow, liver and kidney function defined as
   - Blood white blood cell ≥ lower limit of normal
   - Blood neutrophil count ≥ $1x10^9$/L
   - Blood platelet count ≥ $100x10^9$/L, for HCC ≥ $50x10^9$/L
   - Blood haemoglobin ≥ 9.0 g/dL
   - Creatinine clearance > 40 mL/min calculated by Cockcroft-Gault formula

- AST ≤ 3 X ULN (≤ 5 x ULN when HCC or hepatic metastases are present)
- ALT ≤ 3 X ULN (≤ 5 x ULN when HCC or hepatic metastases present)
- Bilirubin ≤ 1.5 X ULN
- Albumin ≥ 3.0 g/dL

9. Women of child-bearing potential must have a negative pregnancy test in serum prior to trial entry
10. Women of child-bearing potential and men who have partners of child-bearing potential must be willing to practise highly effective contraception for the duration of the trial and for three months after the completion of treatment

**Exclusion Criteria**

1. Less than 21 days since the last dose of intravenous anticancer chemotherapy or less than five half-lives from a small molecule targeted therapy or oral anticancer chemotherapy before the first IMP administration
2. Any immunotherapy within preceding 6 weeks from the first IMP administration
3. Investigational therapy or major surgery within 4 weeks before the first IMP administration
4. Active clinically serious infection > Grade 2 NCI-CTCAE version 5.0 (Appendix 5 - Common Toxicity Criteria Gradings) within preceding 2 weeks before the first IMP administration
5. Brain metastases
6. Subject has not recovered from the previous therapies to Grade ≤ 1 severity as classified by the NCI-CTCAE version 5.0 (except Grade ≤ 2 alopecia, neuropathy or thyroid disorders)
7. Pregnant or lactating women
8. History of second malignancy except for non-melanotic skin cancer, cervical carcinoma in situ or superficial bladder cancer, or any other malignancy treated previously with curative intent and more than three years without relapse
9. Evidence of severe or uncontrolled systemic diseases, congestive cardiac failure New York Heart Association (NYHA) class ≥ 2 (Appendix 7 - NYHA classification), Myocardial Infarction (MI) within 6 months or laboratory finding that in the view of the investigator makes it undesirable for the subject to participate in the trial
10. Any medical condition that the Investigator considers significant to compromise the safety of the subject or that impairs the interpretation of IMP toxicity assessment

11. Confirmed human immunodeficiency virus infection
12. Symptomatic cytomegalovirus infection
13. Subjects with active auto-immune disorder (except type I diabetes, celiac disease, hypothyroidism requiring only hormone replacement, vitiligo, psoriasis, or alopecia)
14. The subject requires systemic corticosteroid or other immunosuppressive treatment
15. Subjects with organ transplants
16. Subjects in dialysis
17. Use of Live (attenuated) vaccines for 30 days prior to the start of study treatment, during treatment, and until last visit
18. Subject is unwilling or unable to comply with treatment and trial instructions
19. Subjects with known hypersensitivity to the IMP or any of the pharmaceutical ingredients

**Specific Additional Exclusion Criteria for HCC**

1. Any ablative therapy (Radio Frequency Ablation or Percutaneous Ethanol Injection) for HCC (this should not exclude subjects if target lesion(s) have not been treated and occurred > 6 weeks prior trial entry
2. Hepatic encephalopathy
3. Ascites refractory to diuretic therapy
4. Child-Pugh score ≥ 7

#### Table S2: Treatment related adverse events part I and part II separated by dose

| Dose (mg/kg) | 0.1 | 0.3 | 1.0 | 3.0 | 10 | Total |
|---|---|---|---|---|---|---|
| | (n = 5) | (n = 13) | (n = 97) | (n = 17) | (n = 6) | (n = 138) |
| | n (%) | n (%) | n (%) | n (%) | n (%) | n (%) |
| **Any** | 3 (60.0) | 8 (61.5) | 49 (50.5) | 6 (35.3) | 3 (50.0) | 69 (50.0) |
| **Blood and lymphatic system disorders** | | | | | | |
| Anaemia | 1 (20.0) | 0 (0.0) | 6 (6.2) | 1 (5.9) | 0 (0.0) | 8 (5.8) |
| **Gastrointestinal disorders** | | | | | | |
| Nausea | 1 (20.0) | 1 (7.7) | 5 (5.2) | 0 (0.0) | 0 (0.0) | 7 (5.1) |
| Vomiting | 0 (0.0) | 0 (0.0) | 4 (4.1) | 0 (0.0) | 0 (0.0) | 4 (2.9) |
| **General disorders** | | | | | | |
| Fatigue | 1 (20.0) | 5 (38.5) | 14 (14.4) | 2 (11.8) | 1 (16.7) | 23 (16.7) |
| Pyrexia | 1 (20.0) | 2 (15.4) | 6 (6.2) | 2 (11.8) | 1 (16.7) | 12 (8.7) |
| **Investigations** | | | | | | |
| Blood alkaline phosphatase increased | 0 (0.0) | 0 (0.0) | 5 (5.2) | 2 (11.8) | 0 (0.0) | 7 (5.1) |
| Alanine aminotransferase increased | 0 (0.0) | 0 (0.0) | 3 (3.1) | 1 (5.9) | 0 (0.0) | 4 (2.9) |
| Aspartate aminotransferase increased | 0 (0.0) | 0 (0.0) | 2 (2.1) | 2 (11.8) | 0 (0.0) | 4 (2.9) |
| **Metabolism and nutrition disorders** | | | | | | |
| Decreased appetite | 0 (0.0) | 0 (0.0) | 3 (3.1) | 1 (5.9) | 0 (0.0) | 4 (2.9) |

*Included are treatment related adverse events with National Cancer Institute-Common Terminology Criteria for Adverse Events (NCI-CTCAE) version 5.0 that occurred in at least in four patients.

#### Table S3: Treatment emergent adverse events part I and part II

| Treament emergent adverse event* | | All grades (n=138) | Grade ≥3 (n=138) |
|---|---|---|---|
| | | number (percent) | |
| **Any** | | 138 (100.0) | 111 (80.4) |
| **Blood and lymphatic system disorders** | | | |
| | Anemia | 32 (23.2) | 8 (5.8) |
| **Gastrointestinal disorders** | | | |
| | Abdominal pain | 33 (23.9) | 4 (2.9) |
| | Constipation | 24 (17.4) | 1 (0.7) |
| | Nausea | 21 (15.2) | 0 |
| | Diarrhoea | 16 (11.6) | 0 |
| | Vomiting | 15 (10.9) | 1 (0.7) |
| | Ascites | 13 (9.4) | 8 (5.8) |
| | Intestinal obstruction | 3 (3.1) | 3 (2.2) |
| | Small intestinal obstruction | 2 (1.4) | 2 (1.4) |
| | Large intestinal obstruction | 2 (1.4) | 2 (1.4) |
| | Ileus | 2 (1.4) | 2 (1.4) |
| **General disorders and administration site conditions** | | | |
| | Fatigue | 51 (37.0) | 4 (2.9) |
| | Pyrexia | 20 (14.5) | 0 |
| | Oedema peripheral | 7 (5.1) | 0 |
| | Death | 7 (5.1) | 7 (5.1) |
| | General physical health deterioration | 4 (2.9) | 3 (2.2) |
| **Hepatobiliary disorders** | | | |
| | Cholestasis | 6 (4.3) | 0 |
| | Hepatic failure | 2 (1.4) | 2 (1.4) |
| **Infections and infestations** | | | |
| | Pneumonia | 3 (2.2) | 2 (1.4) |
| **Investigations** | | | |
| | Blood alkaline phosphatase increased | 21 (15.2) | 3 (2.2) |
| | Aspartate aminotransferase increased | 17 (12.3) | 3 (2.2) |
| | Alanine aminotransferase increased | 16 (11.6) | 1 (0.7) |
| | Blood bilirubin increased | 8 (5.8) | 3 (2.2) |
| | Transaminases increased | 3 (2.2) | 3 (2.2) |
| **Metabolism and nutrition disorders** | | | |
| | Decreased appetite | 24 (17.4) | 0 |
| | Hyperglycaemia | 6 (4.3) | 3 (2.2) |
| | Hyponatraemia | 5 (3.6) | 2 (1.4) |
| **Musculoskeletal and connective tissue disorders** | | | |
| | Back pain | 13 (9.4) | 2 (1.4) |
| | Flank pain | 9 (6.5) | 0 |
| | Myalgia | 6 (4.3) | 0 |
| | Pain in extremity | 5 (3.6) | 0 |
| | Arthralgia | 5 (3.6) | 0 |
| **Neoplasms benign, malignant, and unspecified** | | | |
| | Malignant neoplasm progression | 124 (89.9) | 88 (63.8) |
| | Metastases to central nervous system | 4 (2.9) | 3 (2.2) |
| | Tumor pain | 4 (2.9) | 2 (1.4) |
| **Nervous system disorders** | | | |
| | Headache | 8 (5.8) | 0 |
| | Dizziness | 5 (3.6) | 0 |
| **Psychiatric disorders** | | | |
| | Insomnia | 7 (5.1) | 0 |
| **Respiratory, thoracic and mediastinal disorders** | | | |
| | Dyspnoea | 13 (9.4) | 1 (0.7) |
| | Cough | 6 (4.3) | 0 |
| | Pleural effusion | 4 (2.9) | 3 (2.2) |
| **Vascular disorders** | | | |
| | Hypertension | 4 (2.9) | 2 (1.4) |

*Included are treatment emergent adverse events that occurred in at least five patients or treatment emergent adverse grade ≥3 adverse events that occurred in at least two patient.

Table S4: PFS on bexmarilimab/duration of previous treatment line ratio of > 1.3

| | | n (%) | p-value* |
|---|---|---|---|
| All | | 130 (100) | |
| | Yes | 21 (16) | |
| | No | 109 (84) | |
| Non-DC | | 111 (100) | |
| | Yes | 13 (12) | |
| | No | 99 (88) | |
| DC | | 19 (100) | |
| | Yes | 8 (42) | 0.0031 |
| | No | 11 (58) | |

*Two sided Fisher's exact test

#### The analysis population sets

1. DLT population (n=30): all the patients treated in part I dose escalation cohorts who have received at least one dose of bexmarilimab and have at least three-week follow-up period after the 1st dose.
2. Safety population (n=138): all the patients who had received at least one dose of bexmarilimab
3. Efficacy analysis populations (ORR, PFS, OS, duration of response) (n=138): all the patients who had received at least one dose of bexmarilimab
4. Waterfall blot analysis (n=23): all the patients who had received at least one dose of bexmarilimab and had data from the follow-up RECIST 1.1 evaluation available
5. Landmark analysis set (n=91): all the study patients who are alive at cycle four time-point (9 weeks after 1st dose of bexmarilimab)
6. Previous line therapy duration population (n=134): all the study subject who had received at least one dose of bexmarilimab, had received previous line therapy for advanced disease, had accurate dates available for the beginning and end of the treatment
7. Bexmarilimab pharmacokinetics population (n=30): All the study patients treated in part I and had repeated pharmacokinetic samples available
8. Clever-1 receptor occupancy population (n=28): All the study subject treated in part I of the trial and had samples available for RO analysis
9. sClever-1 populations (n=28): All the study subject treated in part I of the trial and had repeated samples available for sClever-1 analysis
10. Tumor Clever-1 and PD-L1 IHC populations (n=78/43): all the study subject who had received at least one dose of bexmarilimab, belonged to the specific cohorts with high DC rates ≥25% (cutaneous melanoma, gastric, BTC, and HCC), who had adequate pre-treatment tumor samples available for analysis, and had successful IHC staining and positive cell scoring results available.

#### Experimental methods for PK, RO, s-Clever, and tumor biomarker analysis

**PK analysis**

Anti-bexmarilimab Fab Fragment AbD30055 (Bio-Rad) was coated on 96-well plates and blocked. Dilution series of bexmarilimab was used for standard curve preparation. The standard and the diluted samples were added to the wells. After washing, the assay was visualized by the subsequent additions of HRP-labelled mouse anti-human IgG4 (Fc) antibody and a chromogenic substrate (TMB). The concentration of bexmarilimab in samples was back calculated from a calibration curve.

**RO analysis**

PBMCs were plated at 0.2×106 cells/well in round-bottom 96-well plates. All wells were stained with anti-human CD14-Pacific Blue (clone M5E2, BD Pharmingen) together with 10ng/µl in-house conjugated (AF647) anti-Clever-1 antibodies 9-11 or FP-1305. Irrelevant isotype control antibodies ratIgG2a or human in-house conjugated IgG4 (S241/L248E) were used for signal normalization, respectively. FACS was run on the LSRFortessa (BD) and analyzed with FlowJo software v. 10.7.1 (TreeStar).

**sClever-1 analysis**

Serum samples were analyzed for sClever-1 expression using a sandwich-ELISA (enzyme-linked immunosorbent assay) method in which capture antibodies (9-11), blocking agents, samples, detecting antibodies (biotinylated bexmarilimab), and finally detecting agents are introduced to the wells in successive incubation periods followed by a washing step in between. The concentration is determined by comparing the amount of Europium-labelled antibodies in the samples to the Europium-labelled antibodies in the reference standard of single donor lymph.

**IHC analysis for Clever-1 and PD-L1**

FFPE samples with 4-5 µm sections were stained with Ventana Benchmark Ultra (Roche Diagnostics, Basel, Switzerland). For Clever-1 stainings, UltraView Universal DAB Detection Kit (Roche Diagnostics) combined with Clever-1 primary antibody (clone 4G9, sc-293254, Santa Cruz, Dallas, TX, USA) at 1:100 dilution was used. PD-L1 staining was performed using 22C3 pharmDx assay (Agilent Technologies, Santa Clara, CA, USA) according to manufacturer's instructions. Interpretation and scoring were performed by board-certified pathologists using bright field microscopy.

Percentage of Clever-1 positive viable cells (the number of all Stabilin-1 positive cells divided by the total number of cells, multiplied by 100) were scored irrespectively of location, intratumorally, and in stroma. PD-L1 was scored as combined positive score (CPS) by calculation the number of PD-L1 staining cells (tumor cells, lymphocytes, macrophages) divided by the total number of viable tumor cells, multiplied by 100.

**Circulating cytokine analysis**

Samples for circulating cytokines were collected on cycles 1-4. On the cycles 1,2, and 4, analysis was done predose, d2, d8, and d15, and on cycle 3, predose. Cytokines were measured from serum with V-PLEX Chemokine Panel 1 Human and V-PLEX Plus Chemokine Panel 1 Human from Meso Scale Discovery (MSD, Rockville, Maryland, US) by Translational Biomarker Solutions, Labcorp Drug Development, UK.
